# Morphology-based classification of sickle cell disease and β-thalassemia using a low-cost automated microscope and machine learning

**DOI:** 10.1101/2024.09.21.24314128

**Authors:** Pranav Shrestha, Hendrik Lohse, Christopher Bhatla, Heather McCartney, Alaa Alzaki, Navdeep Sandhu, Pradip Kumar Oli, Hongquan Li, Manu Prakash, Ali Amid, Rodrigo Onell, Nicholas Au, Hayley Merkeley, Videsh Kapoor, Rajan Pande, Boris Stoeber

## Abstract

Sickle cell disease (SCD) and β-thalassemia are the most common monogenic diseases, disproportionately affecting low- and middle-income countries, where low-cost and accurate diagnostic tools are needed to reduce the global disease burden. Although the sickling test is commonly used to screen for the sickle mutation, it cannot distinguish between the asymptomatic sickle cell trait (SCT) and SCD, or identify β-thalassemia. Here, we enhanced the inexpensive sickling test using automated microscopy and morphology-based machine learning classification to detect SCD, trait conditions (SCT and β-thalassemia trait) and normal individuals with an overall area under receiver operating curve, sensitivity and specificity of 0.940 (95% confidence intervals: 0.938-0.942), 84.6% (84.2%-84.9%), and 92.3% (92.1%-92.4%), respectively. Notably, the sensitivity and specificity to detect severe disease (SCD) was over 97% and 98%, respectively, thus establishing a low-cost automated screening option for disease detection in low-resource settings. Furthermore, leveraging high-throughput microscopy, we generated an open-access dataset comprising over 300,000 images with 1.5 trillion segmented cells from 138 individuals in Canada and Nepal including individuals with sickle and/or β-thalassemia mutations, to accelerate further research.

## Introduction

Sickle cell disease (SCD) is linked with an estimated 300,000-400,000 births every year and a mortality rate exceeding 50% in certain areas of low-income countries^1,2^. SCD is caused by a mutation in the gene encoding hemoglobin β-globin subunit (HBB) resulting in a variant form of hemoglobin (HbS or sickle hemoglobin). HbS causes red blood cells (RBCs) to distort into rigid “sickled” cells that can lead to blockage of blood vessels (vaso-occlusion)^1,3,4^, hemolysis, inflammation and progressive organ damage^4,5^. β-thalassemia is another inherited disease most commonly caused by point mutations in HBB, resulting in either a reduced (β^+^) or absent (β^0^) synthesis of β chains of hemoglobin^6^. SCD is an umbrella term for a combination of inherited diseases, including sickle cell anemia (homozygous HbSS) and compound heterozygous sickle cell disease (HbS/β-thalassemia, HbS/C, *etc.*). Inheritance of a mutated gene from only one parent results in clinically asymptomatic, but carrier forms, such as sickle cell trait (SCT; HbAS) and β-thalassemia trait (HbA/β-thalassemia)^7^, which are usually more prevalent than disease forms^5^. Screening for trait conditions (SCT and β-thalassemia trait) is also essential to prevent the variant genes from being unknowingly transferred to children, potentially even as severe disease forms.

Globally, HbS has the highest prevalence in sub-Saharan Africa, while HbS and β-thalassemia are both prevalent in the Middle East, Mediterranean countries and South Asia^4–6^. Although the genetics and hemoglobin biophysics of SCD, the first “molecular disease”^8^, have been extensively studied^4^, and promising new techniques such as CRISPR-Cas9 gene editing for SCD and β-thalassemia have been clinically tested^9^, there is still an unmet need to overcome the diagnostic barrier in low-resource settings, where the burden of the disease is disproportionately high^10,11^. Access to screening, caregiver education and treatment options greatly improve quality of life and life expectancy of people with SCD, and reduce childhood mortality by up to 90%^1,12^. However, the burden of the disease is highest in resource-constrained regions of low- and middle-income countries, with limited access to such screening and treatment options. To enable timely and equitable diagnosis, low-cost and point-of-care detection techniques are required in such low-resource settings to supplement or replace expensive clinical laboratory techniques^11^.

Gold standard tests for detecting most hemoglobinopathies like SCD and β-thalassemia include hemoglobin (Hb) electrophoresis and high performance liquid chromatography (HPLC), which rely on separating different variants of hemoglobin based on charge^13^. Hb HPLC is run on expensive equipment in clinical laboratories by highly trained personnel, thus is not suitable for disease detection in rural/remote or resource-constrained settings. Conventional low-cost alternatives, such as the sickling test and Hb solubility test, detect the presence of HbS, but do not differentiate between SCT and SCD. In the sickling test, blood is sealed between a microscope slide and a coverslip with a reagent that causes deoxygenation (hypoxia), which induces sickling of RBCs for individuals with SCT and SCD^7,13^. The solubility test relies on the principle that HbS is insoluble in a high molarity phosphate buffer when deoxygenated, resulting in a turbid solution (positive test for SCT and SCD)^7^.

Recently, novel approaches for detecting SCD have been developed or commercialized. Two commercially available lateral-flow assays, HemoTypeSC and Sickle SCAN, detect HbS and variant hemoglobin C (HbC), with reported sensitivity and specificity of over 92%^14–20^. A low-cost paper-based electrophoresis platform, the Gazelle Hb variant test, has also been commercialized to detect HbS, HbC and β-thalassemia^21^. The lateral flow assays and the Gazelle cost between $2 to $5 per test (with an additional one-time cost of $1200 for the Gazelle portable reader), and are around an order of magnitude costlier per test than the sickling test. Despite the importance of screening for β-thalassemia (including β-thalassemia trait) in areas with high prevalence of both HbS and β-thalassemia, none of the commercialized low-cost techniques (except Gazelle) detect β-thalassemia.

The morphological differences between sickled and round RBCs can be observed through microscopy, which makes digital microscopy, augmented with machine learning, a potentially powerful tool for disease identification. Most machine learning approaches thus far focus on identifying sickle cells in microscopy images of peripheral blood smears, using techniques such as semantic segmentation^22^, or classification on segmented cells from datasets using lightweight models (*e.g.* support vector machine multi-class classifiers)^23^ or deep convolutional neural networks^24^. Although high accuracy of 98-99% has been demonstrated using such techniques, sickle cells are usually only observed in peripheral blood smears for SCD samples and not for SCT samples, thus preventing the use of such techniques to screen for trait conditions. To overcome this limitation, another approach analysed microscopy images of the sickling test, instead of peripheral blood smears. SCD and SCT were distinguished based on morphological differences between cells using high and low concentrations of a sickling reagent, but this technique required using two reagent concentrations and a custom microfluidic chamber^25^.

Furthermore, most of these machine-learning based techniques have only been validated on thousands of cells, thus highlighting a key challenge with these techniques - *i.e.* having limited annotated data for training models. Although transfer learning, data augmentation and lightweight model architectures can be used to work with small datasets^23^, high-throughput microscopy can potentially generate large image datasets and aggregation of morphological data can be utilized to avoid annotating at the single cell level (when patient-level ground truth is available).

Here, we introduce an augmented version of the inexpensive sickling test using low-cost automated microscopy and machine learning to detect SCD, SCT and β-thalassemia (Fig. 1a). We address the main shortcomings of the sickling test (inability to distinguish between SCT and SCD, inability to detect β-thalassemia, and requirement of trained personnel to find sickle cells under a microscope), while retaining its strengths (cost-effectiveness, basic sample preparation using one concentration of the sickling reagent and easily available consumables, no requirement for staining or fixing cells, and reliance on the morphology of RBCs for detection). Using high-throughput low-cost microscopy, we generated a large open-access dataset of microscopic images of blood cells from de-identified samples collected at clinical sites in Canada and Nepal, suitable for statistical analysis and machine learning. Our classification approach is based on morphological differences between groups, relying on differences in cell shapes between normal and sickled cells, differences in the level of sickling between SCD and SCT, and differences in cell shape and size between normal and β-thalassemia trait. The study aims to enable inexpensive and automated screening of SCD and trait conditions (SCT, β-thalassemia trait), and improve access to screening in low-resource settings.

**Fig. 1.**
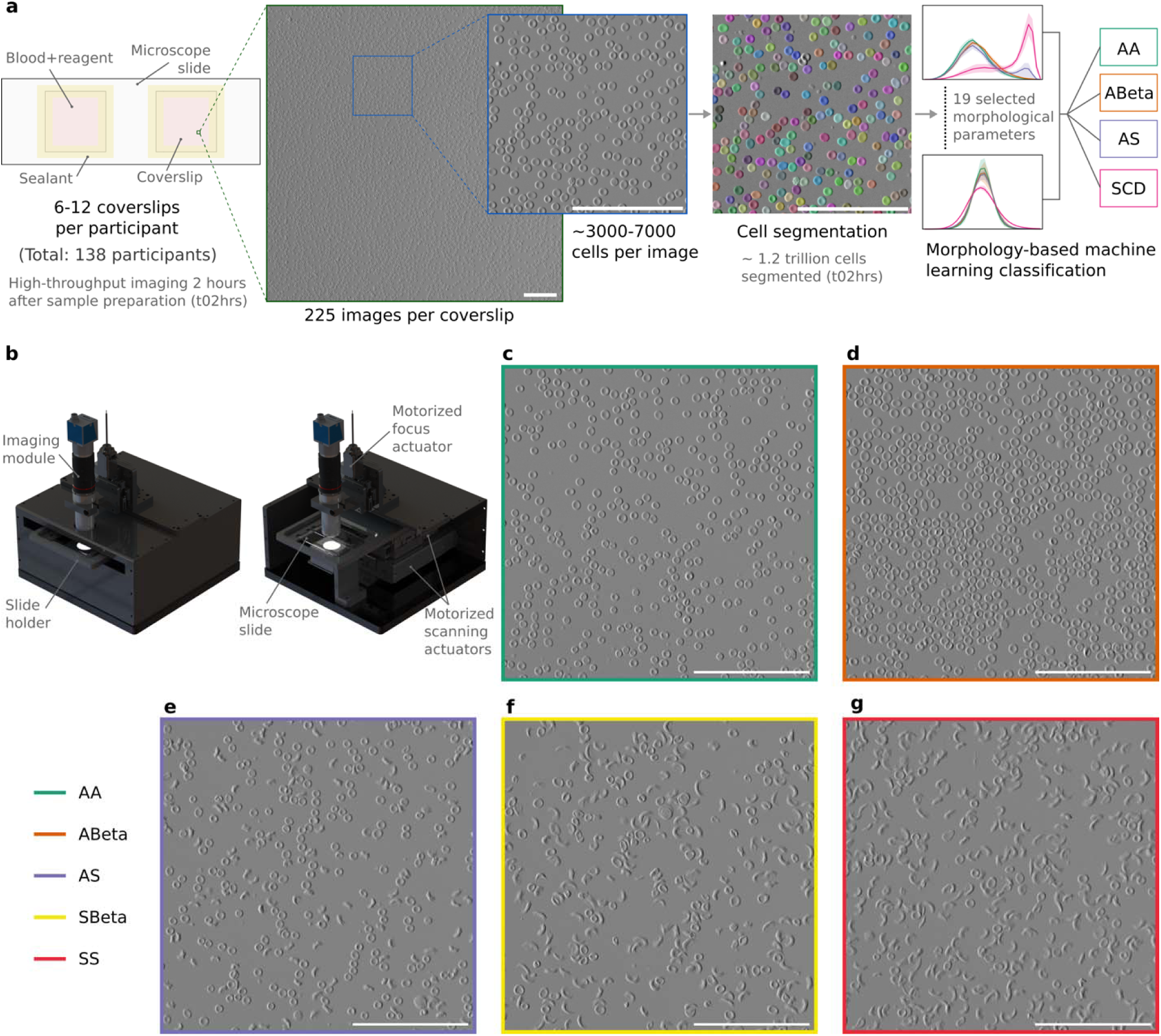
High-throughput imaging for morphology-based classification. **a**, Sequence of sample preparation, imaging, cell segmentation, and classification. **b**, Computer-aided design (CAD) renderings of automated microscope, Octopi, with enclosure (left) and without some enclosure panels (right), showing different components. Picture of microscope during operation in Fig. S1 in Supplementary information. **c-g**, Representative image segments from normal (AA = HbAA), β-thalassemia trait (ABeta = HbA/β-thalassemia), sickle cell trait (AS = HbAS), compound heterozygous sickle cell disease (SBeta = HbS/β-thalassemia), and homozygous sickle cell disease (SS = HbSS). Each image segment is 270 µm × 270 µm, cropped from 900 µm × 900 µm images (original FOVs). Images were taken two hours after sample preparation for the sickling test (blood + 2% sodium metabisulphite in 1:2 volume ratio). White scale bars in images (**a**, **c**-**g**) are 100 µm.

## Results

### High-throughput automated microscopy of sickling test and creation of open-access image dataset

A total of 138 participants were recruited in Nepal and Canada: 30 without β-globin disorders (HbAA), 23 with β-thalassemia trait (HbA/β-thalassemia), 45 with SCT (HbAS), and 40 with SCD (11 HbS/β-thalassemia and 29 HbSS). We modified the sample preparation protocol of the conventional sickling test for high-throughput imaging and cell segmentation (more details in Methods). For each participant, 3-6 microscope slides (with 2 coverslips each) were prepared (Fig. 1a) and 225 images were captured per coverslip in around 4 minutes by the Octopi microscope^26,27^, with automated slide scanning and automated focusing (Fig. 1b). Two hours after sample preparation, RBCs from normal (HbAA; Fig. 1c) and β-thalassemia trait (HbA/β-thalassemia; Fig. 1d) participants appeared unchanged to the eye, while hypoxia-induced sickling was observed (Fig. 1e-g) for the majority of RBCs from SCD participants (HbS/β-thalassemia and HbSS), and for a fraction of RBCs from SCT participants (HbAS). The process of sickling, facilitated by the polymerization of HbS, depends on factors such as HbS concentration, temperature and rate of deoxygenation^28,29^. We followed consistent sample preparation and imaging protocols, such that major differences between sickling of different samples resulted from differences in HbS concentration, *i.e.* between SCT and SCD. Imaging 1001 coverslips from 138 participants, around 220,000 images were captured 2 hours after sample preparation and around 1.2 trillion cells were segmented using Cellpose 2.0^30^ for statistical analysis and morphology-based machine learning classification. All the de-identified microscopic images (raw and processed), cell segmentation data, and calculated morphological parameters for images captured at 2 hours and other parameter settings (*e.g.* other time instances, other temperature configurations, time-series data) are publicly available in an open-access database^31^.

### Sickling under hypoxia

The reagent in the sickling test (sodium metabisulphite) and the sealed wet preparation produces an environment of reduced oxygen or hypoxia, which induces sickling in RBCs with HbS (*i.e.* for SCT and SCD). The resolution of imaging (pixel size of 1.85μm for monochromatic camera) using 20× objective microscopy was sufficient to observe sickling at the cellular level (Fig. 2a and Supplementary Videos 1 and 2). Sickling of RBCs produced detectable changes in some morphological parameters over time, even at the level of a single image, containing aggregated data from around 3000 cells (Fig. 2b-g and Supplementary Video 3). As an example for a participant with SCD, within a 50 minute time period, the distribution of area remained nearly constant (Fig. 2b), the perimeter distribution changed slightly (Fig. 2c), and the distributions of major axis, minor axis, roundness, and eccentricity changed considerably (Fig. 2d-g). The time evolution of the morphological parameters reflected the dynamics of RBC sickling, *e.g.* the roundness peak (Fig. 2f) shifted from closer to 1 (round) to around 0.5 (elongated or sickled), and the eccentricity peak (Fig. 2g) shifted from around 0.4 (less elongated) to around 0.9 (more elongated or sickled), indicating that greater proportions of cells sickled over time.

**Fig. 2.**
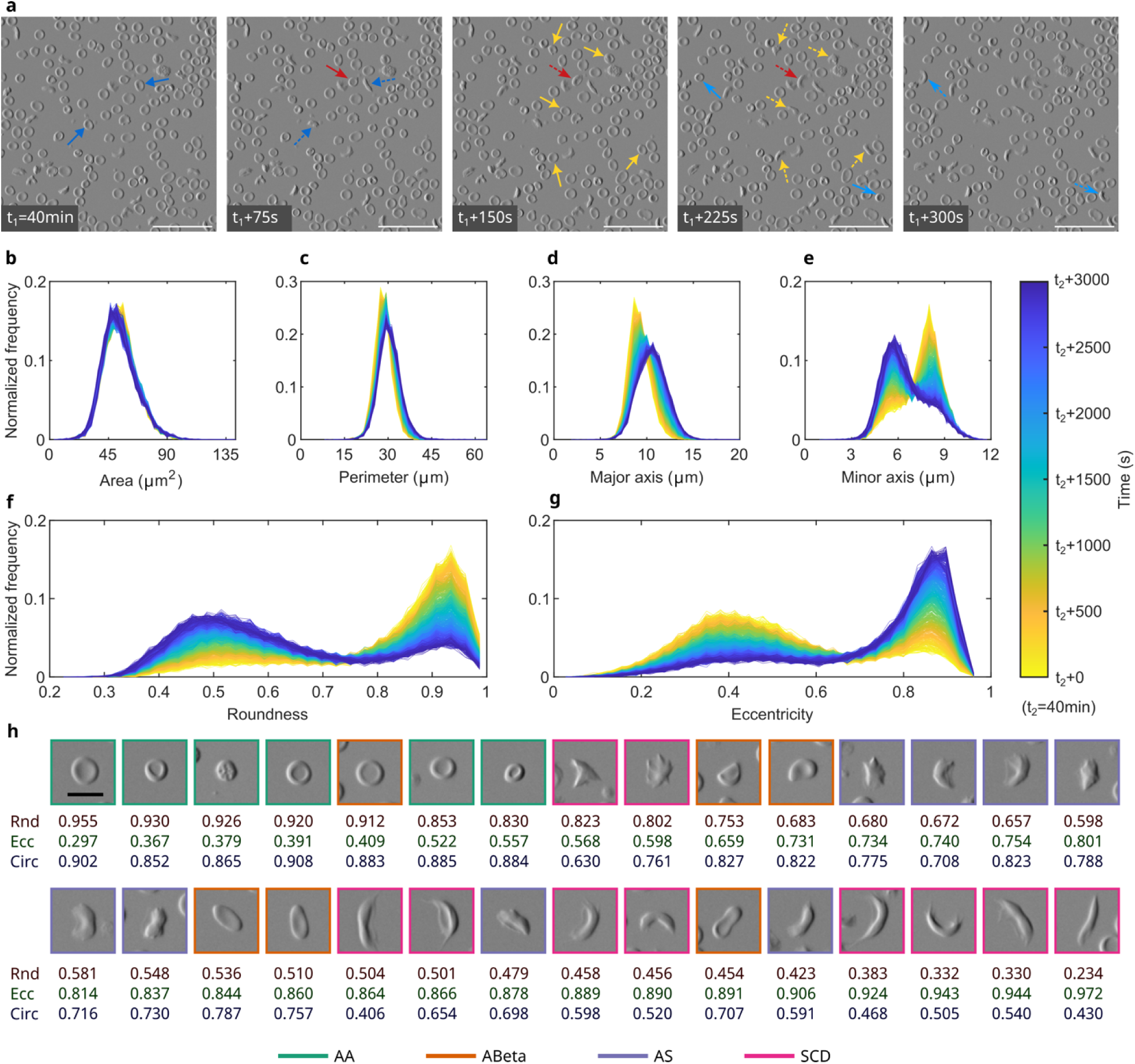
Morphological changes during sickling and examples of cells from different groups. **a**, Time series of image segments for a sample from a sickle cell disease (HbSS) participant, showing sickling at the cellular level; Solid and dashed arrows show cells before and after sickling, respectively; Time interval between adjacent images is 75 seconds; Scale bars (white) are 50 µm. **b-g**, Normalized frequency over time of different morphological parameters for an image (900 µm x 900 µm); Time series includes 1000 instances (or curves of frequency distribution), for a total time period of 3000 seconds; Number of cells = 2805 ± 17 (mean ± standard deviation for 1000 instances); Morphological parameters changed between initial (yellow) and final (blue) time instances with varying degrees for area (**b**), perimeter (**c**), major axis (**d**), minor axis (**e**), roundness (**f**), and eccentricity (**g**). Supplementary video 3 shows the sickling of RBCs over time at the image level at 120× speed. **h**, Examples of cells with different values of morphological parameters, arranged in descending order of roundness (Rnd), ascending order of eccentricity (Ecc), and not sorted for circularity (Circ); Scale bar (black) in the first image is 10 µm.

### Morphological characterization at multiple scales

Blood cells in microscopic images were segmented by fine-tuning a neural network-based model (Cyto) in Cellpose 2.0 using a human-in-the-loop approach^30^. Based on the outlines of the segmented cells, 40 different parameters describing the size (*e.g.* area, perimeter), shape (*e.g.* roundness, eccentricity), and intensity (*e.g.* mean intensity) of the cells were calculated (complete list in Supplementary information). Different types of cells, including different variations of sickled cells (*e.g.* crescent, holly leaf, granular), resulted in different values for morphological parameters (Fig. 2h). The morphological parameters determined at the cellular level were aggregated at different levels or scales – group level (colored lines in Fig. 3), donor level (Fig. 3a), slide level, coverslip level (Fig. 3b), and image level (Fig. 3c). For instance, the frequency distribution of eccentricity, which describes how elongated a shape is, was calculated for all the cells per donor or participant (aggregating data for nearly 6-10 million cells per participant; Fig. 3a). The distribution of eccentricity for normal (“AA” or HbAA) and β-thalassemia trait (“ABeta” or HbA/ β-thalassemia) were similar for image, coverslip and donor levels, with peaks close to 0.4 (less elongated). The distribution of eccentricity of SCT (“AS” or HbAS) had two peaks at the group level (colored lines in Fig. 3), indicating that a portion of the cells were sickled (high eccentricity) while the rest were not (low eccentricity). As indicated by the variability of frequency distribution profiles, not all images in the SCT group contained sickled cells. In contrast, the SCD group (*i.e.* HbSS and HbS/β-thalassemia) had prominent peaks at high values of eccentricity at all levels of analysis, indicating that the images contained predominantly sickled or elongated cells. Overall, the curves for SCD were markedly different from the rest, while curves for AA and ABeta, and for AS and the rest overlapped.

**Fig. 3.**
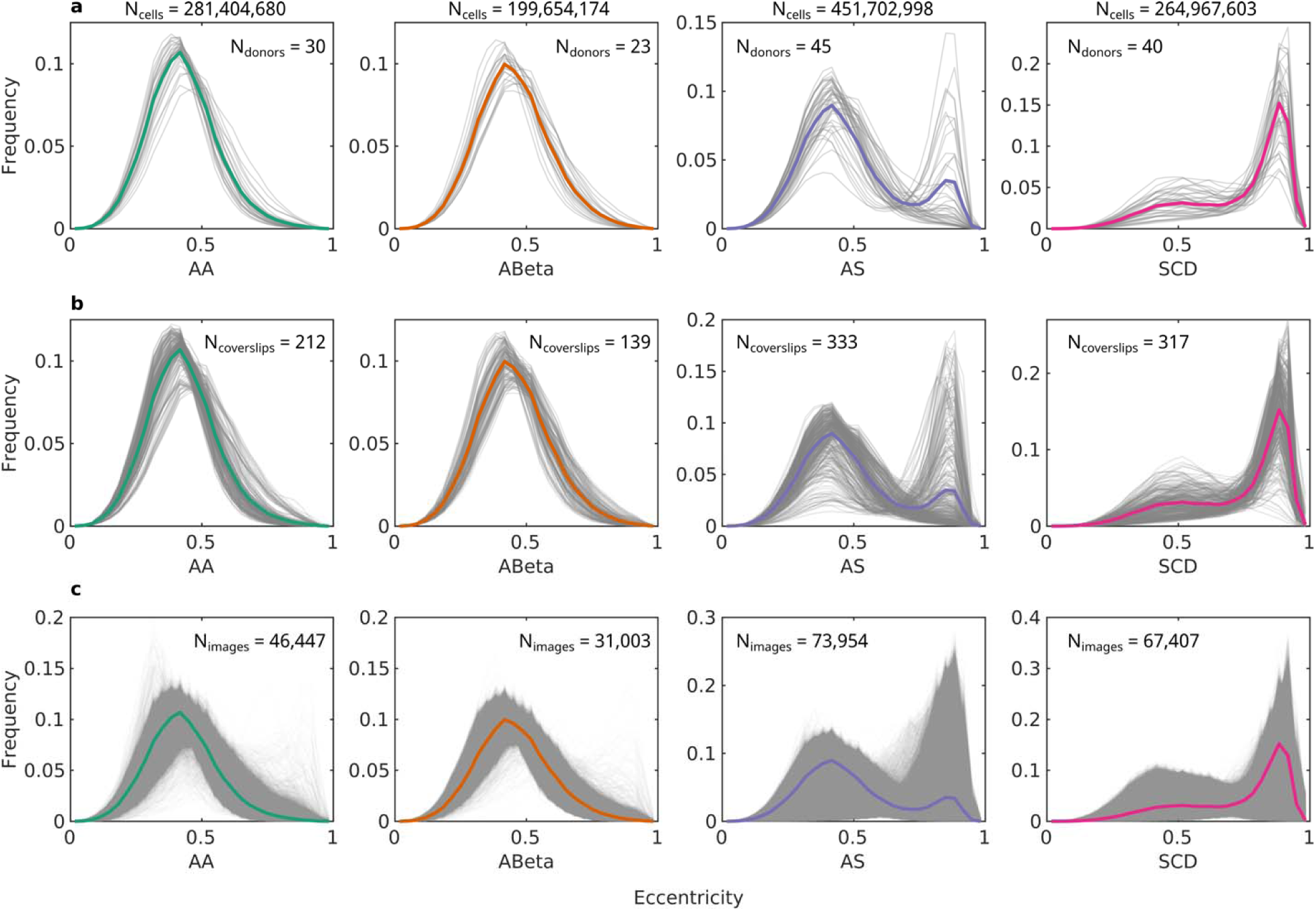
Morphological characterization at different scales. **a**, Normalized frequency distribution of eccentricity for the differen groups (AA, normal; ABeta, β-thalassemia trait; AS, sickle cell trait; SCD, sickle cell disease) at the donor or participant level, where each grey line represents aggregated results for all cells from each participant. **b**, Normalized frequency distribution of eccentricity at the coverslip level, where each grey lines represent aggregated results for all cells in each coverslip. **c**, Normalized frequency distribution of eccentricity at the image level, where each grey line represents aggregated results for all cells in each image. The colored lines (**a**-**c**) represent aggregated results for the entire group. The number of cells (N_cells_) in each group are listed above all plots, while the number of donors (N_donors_), coverslips (N_coverslips_) and images (N_images_) are listed inside the plots.

### Statistical differences between morphological parameters of different groups

To explore whether morphological differences existed between different groups, and to evaluate the feasibility of morphology-based classification, we calculated statistical differences between frequency distributions of different groups for all 40 morphological parameters. This statistical analysis used data for morphological parameters aggregated at the coverslip-level, with a total of 1001 coverslips from 138 participants for 4 clinically relevant groups ‒ AA (HbAA), ABeta (HbA/β-thalassemia), AS (HbAS), and SCD (HbS/β-thalassemia and HbSS). For each morphological parameter, there were parameter ranges (or bins in frequency distribution curves or features for classification) without significant differences (p-value > 0.05) for morphologies of different classes (orange in heatmaps in Fig. 4). In contrast, there were many bins or features that resulted in significant differences (p-value < 0.05) between morphologies of different (or all) groups, thus being important features for morphology-based classification. For example, there were significantly different occurrences of cells with major axis between 10.5 µm and 11 µm for 5 out of 6 combinations (grey box in Fig. 4a; and 4c-d), and with roundness between 0.89 and 0.92 for all 6 combinations (Fig 4e, 4g-h).

**Fig. 4.**
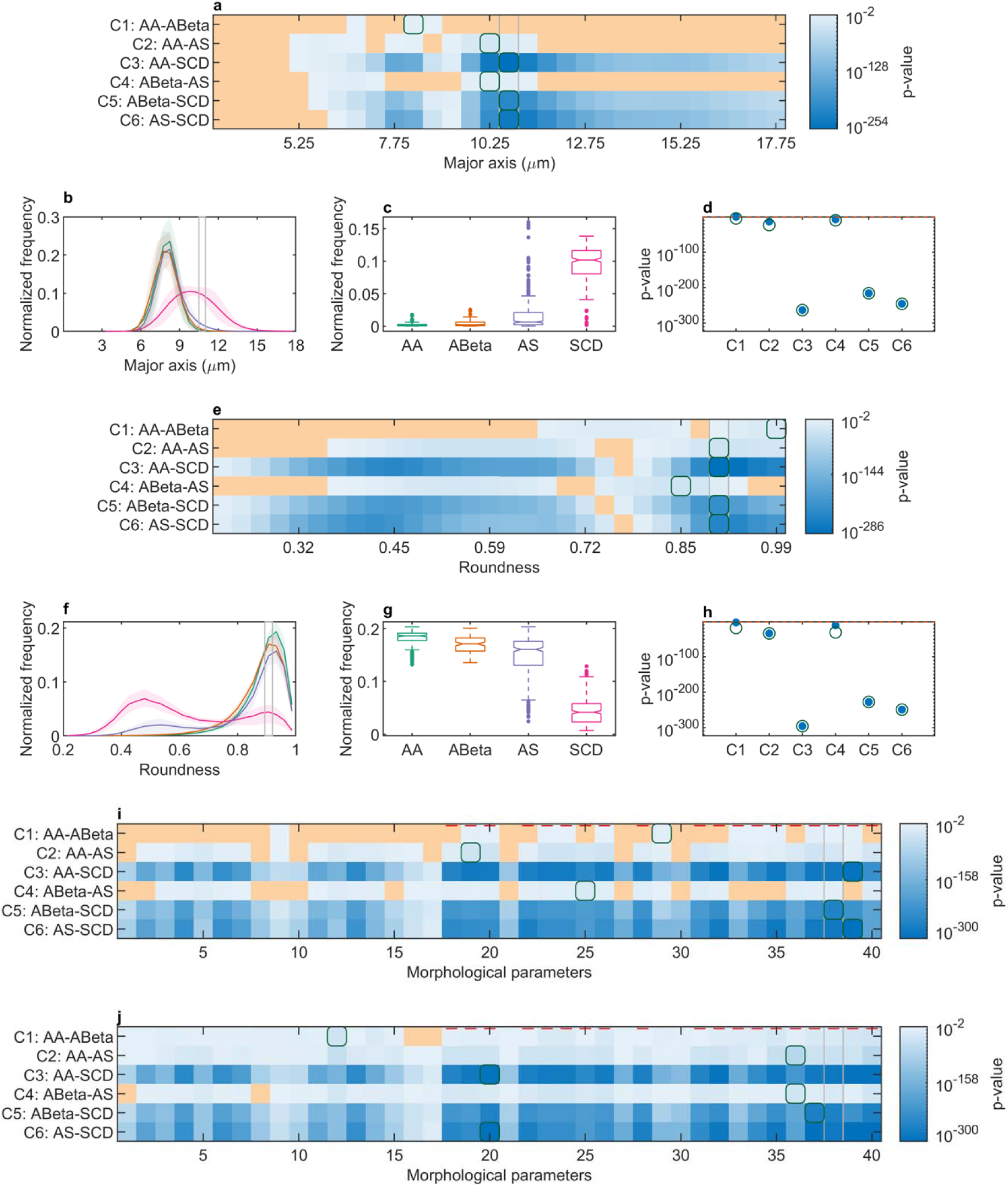
Statistical differences between frequency distributions of different groups at the coverslip level. **a**, p-values for pairwise comparison of means of different groups at each bin using Scheffe’s method, for morphological parameter major axis. **b**, Normalized frequency distribution (30 bins) of major axis for different groups, corresponding to heatmap (in a). **c**, Box plot for major axis between 10.5 µm and 11 µm (highlighted by grey boxes in **a and b**), indicating that there were significantly more cells for SCD with major axis between 10.5 µm and 11 µm than the other groups. d, Minimum overall p-values for all combinations (grey boxes in a and b) shown as blue (filled) circles, and minimum p-values for each combination (green boxes in **a**) shown as green circles. **e-h**, Results for morphological parameter roundness, with grey boxes (in e and f) and data in g and h corresponding to roundness values between 0.89 and 0.92. **i**, Combinations for each morphological parameter with minimum overall (geometric mean) p-values (*e.g.* grey boxes in **a** and **e** appear at the corresponding indices of 4 and 20). **j**, Minimum p-values for each combination of all morphological parameters (*e.g.* green boxes in **a** and **e** appear at the corresponding indices of 4 and 20). Orange in heatmaps (**a**, **e**, **i**, and **j**) indicates p value > 0.05 (difference not statistically significant), green boxes (in heatmaps) and green circles (**d** and **h**) indicate minimum p-values for the respective combination (row), and grey boxes (in heatmaps, and **b** and **f**) indicate minimum overall (geometric mean) p-values for all 6 combinations. Red horizontal lines in heatmaps (**i**, **j**) indicate non-dimensional morphological parameters. Box plots show median (line), 25th and 75th percentiles (box), non-outlier minimum and maximum (whiskers), and outliers (dots) for the normalized frequency in the corresponding bin. Notches that do not overlap have different medians at 5% significance level. A list of the morphological parameters (in **i** and **j**) is provided in Supplementary information.

The statistical analysis results (Fig. 4) relate to observed and expected physical or morphological characteristics of cells. Firstly, the differences between SCD and the other classes were larger than the other combinations (indicated by lower p-values for C3, C5 and C6 than for other combinations in Fig 4a,d,e,h), mainly because the cells from SCD samples were predominantly sickled (indicated by shifted peaks in frequency distribution in Fig. 4b,e), unlike cells from the other groups. However, since AA and ABeta, and AS (to some extent) had round RBCs, there were many bins without significant morphological differences for combinations C1, C2, C4 (Fig. 4a,e). Regardless, even for these 3 combinations, there existed bins with significant morphological differences, likely due to the presence of some sickled cells in AS, and the variation in shape (poikilocytosis) and size (anisocytosis) of RBCs expected in ABeta^32^.

To compare the 40 different morphological parameters with respect to their relevance for sample classification, the top bins or features for each morphological parameter were evaluated (Fig. 4i,j). Considering the minimum p-values for each combination of each morphological parameter (Fig. 4j), almost all the parameters (236/240) had features with significant differences (Fig. 4j). Generally, non-dimensional morphological parameters (red lines in Fig. 4i-j), such as roundness, eccentricity, normalized area, *etc.*, produced more significant differences than other parameters, such as area, perimeter, *etc*.

Additionally, non-dimensional parameters are robust and independent of imaging modalities, and thus were selected for classification. Overall, based on statistical analyses, there were morphological differences between the 4 groups at multiple bins for all 6 combinations, suggesting that morphology-based classification, particularly using frequency distribution from non-dimensional parameters, is feasible.

### Morphology-based machine learning classification

Coverslip-level frequency distribution of 19 non-dimensional morphological parameters were used for machine-learning classification, using 80:20 participant-wise splits of training and testing data, which were iteratively randomized 1000 times to obtain 95% confidence intervals (CI) of performance metrics. Different groups were considered (details in Methods): *3Gp*, *3GpSc*, and *4Gp*. For the different cases, confusion matrices and receiver operating characteristic (ROC) curves of top classifiers are provided in Fig. 5. Performance metrics (class-wise and overall) for the top 5 classifiers in each case are provided in Fig. 6 (more metrics in Tables S1-S3 of Supplementary information).

**Fig. 5.**
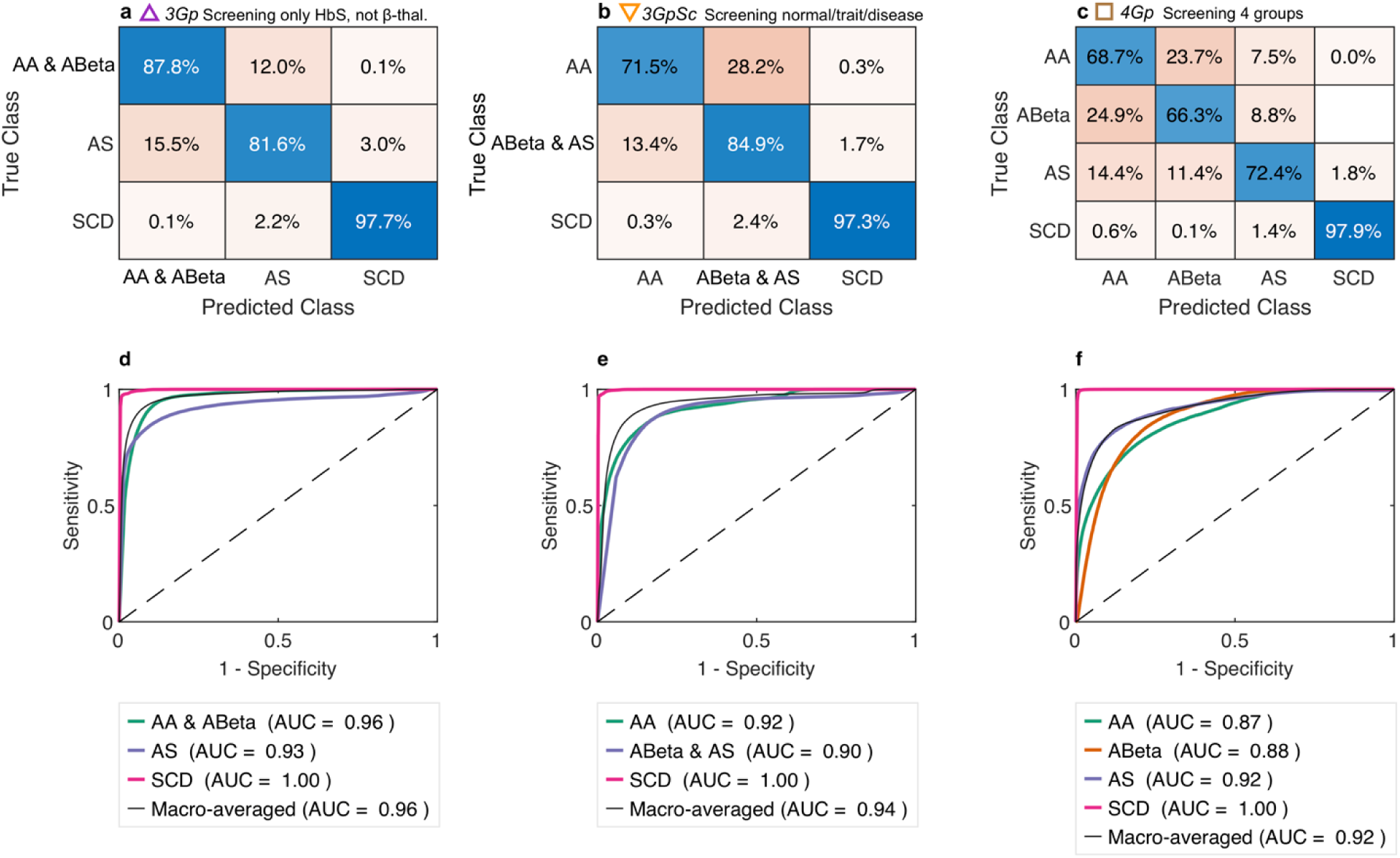
Morphology based classification into different groups. **a**, Confusion matrix for classification into 3 groups (*3Gp*): AA & ABeta, AS, and SCD (SBeta and SS), where diagonal values represent percentages of correct predictions and off-diagonal values represent misclassifications. Classifier: quadratic support vector machine (QSVM). **b**, Confusion matrix for classification into 3 groups suitable for screening both HbS and β-thalassemia (*3GpSc*): AA, trait conditions (ABeta and AS), SCD (SBeta and SS). Classifier: QSVM. **c**, Confusion matrix for classification into 4 groups (*4Gp*): AA, ABeta, AS, SCD (SBeta and SS). Classifier: subspace discriminant (SSD). Confusion matrices (**a**-**c**) are row-normalized. **d-f**, Receiver operating characteristic (ROC) curves for visualizing classifier performance at different thresholds, showing ROC curves for individual classes (one vs. all; colored curves) and macro-averaged ROC curves (black curves) for all groups. Dashed diagonal line (with AUC = 0.5) represents a random guessing classifier that cannot discriminate between positive and negative classes. The areas under the receiver operating characteristic curve (AUC or AUROC) for individual and macro-averaged cases are shown. The confusion matrices and ROC curves show merged results (from 1000 iterations of randomized 80:20 participant-wise splits of training and testing data) for the testing sets only – validation accuracies from training sets were higher likely due to some overfitting, and are not shown. Coverslip-level frequency distribution (30 bins) data for 19 non-dimensional morphological parameters was used for classification.

**Fig. 6.**
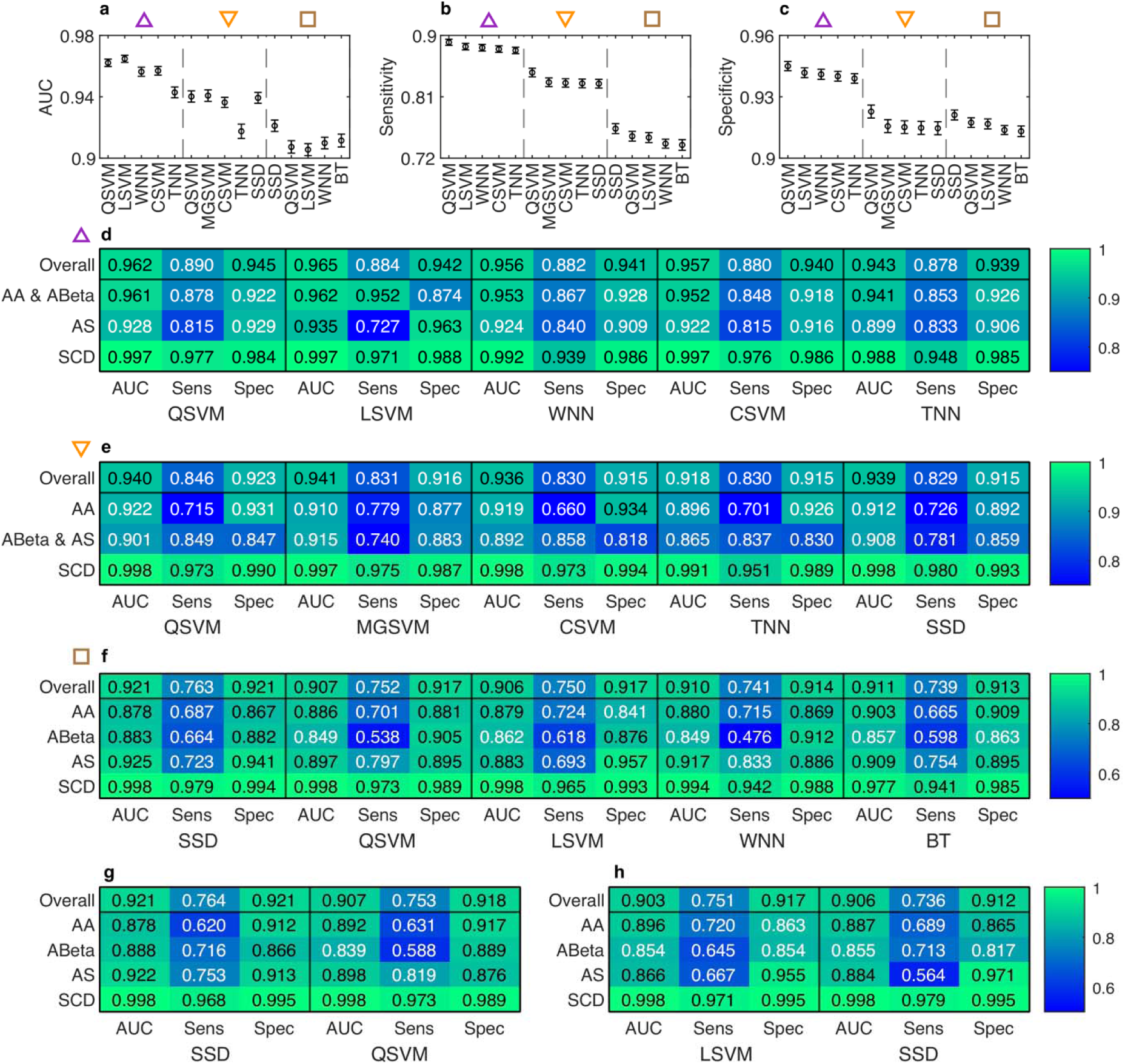
Performance metrics for different classifiers on testing datasets. **a,** Overall macro-averaged AUC (or AUROC) for 5 top classifiers (using 19 non-dimensional morphological parameters at the coverslip-level) to classify into 3 groups (*3Gp*) for screening only for HbS (AA & ABeta, AS, and SCD; purple upright triangle), 3 groups (*3GpSc*) for screening HbS and β-thalassemia (AA, ABeta & AS, and SCD; orange inverted triangle), and 4 groups (*4Gp*; AA, ABeta, AS, SCD; brown square). **b**, Overall macro-averaged sensitivity for the top 5 classifiers of the three cases. **c**, Overall macro-averaged specificity for the top 5 classifiers of the three cases. Error bars (**a**-**c**) represent 95% confidence intervals generated from 1000 iterations of randomized participant-wise splits. **d-f**, Heatmaps showing overall (or macro-averaged) and individual (one vs. all) metrics (AUC, sensitivity and specificity) of the top 5 classifiers for 3 groups (AA & ABeta, AS, and SCD; purple upright triangle; **d**), 3 groups for screening (AA, trait, and SCD; orange inverted triangle; **e**), and 4 groups (AA, ABeta, AS, SCD brown square, **f**). **g**, Heatmaps showing overall and individual metrics of the top 2 classifiers with different misclassification penalties (misclassifications of ABeta, AS, or SCD as normal class AA had misclassification costs 10 times higher than for other misclassifications) **h**, Heatmaps showing overall and individual metrics of the top 2 classifiers with feature selection (77 top features selected from statistical analysis). Abbreviations – AUC, Area under the receiver operating characteristic curve; Sens, Sensitivity; Spec, Specificity; QSVM, Quadratic support vector machine (SVM); LSVM, Linear SVM; WNN, Wide neural network; CSVM, Cubic SVM; TTN, Tri-layered neural network; MGSVM, Medium Gaussian SVM; SSD, Subspace Discriminant; BT, Boosted tree.

The area under the ROC curve (AUC or AUROC) relates to the probability that a classifier will rank a randomly chosen positive case higher than a randomly chosen negative case^33^. For *3Gp*, the top classifier, quadratic support vector machine (QSVM), was able to discriminate the three classes typically considered during HbS screening (Fig. 5a,d, 6d; upright triangles in Fig. 6a-c). The overall or macro-averaged AUC, sensitivity, and specificity were 0.962 (95% CI: 0.961-0.963), 0.890 (0.888-0.892), and 0.945 (0.944-0.946), respectively. Traditionally, the sickling test cannot distinguish between SCT (AS) and SCD, but in this case (*3Gp*), the top machine learning classifier distinguished AA (and ABeta), AS and SCD with class-wise AUC (one vs. all) of 0.961 (0.96-0.963), 0.928 (0.925-0.93) and 0.997 (0.997-0.997), respectively (Fig. 5d, 6d).

For *3GpSc*, the overall AUC, sensitivity and specificity for the top classifier (QSVM) were 0.940 (0.938-0.942), 0.846 (0.842-0.849), and 0.923 (0.921-0.924), respectively (Fig. 5e, 6e, and inverted triangles in Fig. 6a-c). The class-wise AUC for AA, trait (ABeta & AS) and SCD were 0.922 (0.919-0.925), 0.901 (0.898-0.903), and 0.998 (0.998-0.998), respectively. For most of the top 5 classifiers (Fig. 6e), the sensitivity for detecting disease and trait conditions were higher than for AA, indicating that most of the individuals with trait or disease conditions would be identified during screening, and such individuals could be followed up with confirmatory diagnostic tests.

For *4Gp*, the top classifier, subspace discriminant (SSD), was able to discriminate 4 groups (AA, ABeta, AS, SCD) with a macro-averaged AUC of 0.921 (0.919-0.923), sensitivity of 0.763 (0.76-0.767), and specificity of 0.921 (0.92-0.922) (Fig. 5c,f, 6f; squares in Fig. 6a-c). Different misclassification penalties/costs can also be applied depending on the requirements of screening tools, *e.g.* higher penalties for misclassification of trait or disease conditions than normal conditions increased the sensitivity of detecting ABeta and AS (Fig. 6g). Additionally, feature selection can reduce computational resource requirement with minimal effect on classifier performance, *e.g.* selecting the top 77 features based on statistical analysis (Fig. 6h) resulted in comparable classification performance to that using all 570 features from 19 morphological parameters (Fig. 6f).

Notably, the sensitivity for detecting SCD was high (> 97%) in almost all the cases regardless of group divisions, indicating that most of the severe disease cases were detected by the classifiers. This was consistent with results from statistical analysis (Fig. 4) and observations of morphological differences in microscopic images (Fig. 1).

## Discussion

The low-cost sickling test, which is traditionally unable to distinguish between SCT and SCD and unable to detect β-thalassemia, was augmented to detect individuals with/without SCD and trait conditions (including β-thalassemia) with an overall sensitivity and specificity of 84.6% (95% CI: 84.2%-84.9%), and 92.3% (92.1%-92.4%), respectively (with sensitivity to detect only SCD of 97.3%). Such an augmentation of the simple sickling test can enable accurate identification (>97% sensitivity) of people with severe disease who require treatment and medical attention, while also screening for individuals with trait conditions including β-thalassemia (85% sensitivity) who can be informed of the risks of passing variant genes to children. Furthermore, the portable microscope can be implemented in different levels of healthcare facilities without requiring a sophisticated laboratory/clinical setting. Moreover, the same automated microscope can potentially be used for detecting other diseases relying on microscopy, such as malaria^26^ and tuberculosis^34^.

The use of high-throughput automated microscopy for morphology-based classification provides benefits to diagnostics in low-resource settings, allows expert review, and represents a powerful tool for generating large training data sets for the development of advanced classification algorithms. Firstly, automation can be implemented for device operation, slide scanning, and disease detection, reducing the need for highly skilled personnel in low-resource settings. Secondly, if needed, the microscopic images of blood cells can be examined by specialists (*e.g.* hematologists, hematopathologists, laboratory technicians) to validate test results, similar to analysing a conventional sickling test for sickled cells to screen for HbS or a peripheral blood smear for poikilocytosis or anisocytosis of RBCs to screen for β-thalassemia. Furthermore, morphological characterization performed at coverslip-, donor-, or group-levels can aggregate information and statistics of millions to trillions of cells, thus enabling richer analysis at a scale orders of magnitude greater than with individual inspection of images. Such morphological and aggregated characterization of cells relates to physical and observable differences between cells of different groups. Thus, these morphological parameters serve as meaningful inputs to machine-learning based classification and can enhance the interpretability of the classification process and results, which is important for health-care related machine learning applications^35,36^. Lastly, high-throughput imaging enabled the creation of an open-access image dataset with hundreds of thousands of images and trillions of segmented cells with ground truth diagnosis from Hb HPLC, which can be used in the future to further improve classification performance, as segmentation/classification algorithms improve over time. Furthermore, the same dataset can be used for disease detection using other approaches not considered here, such as semantic segmentation or object detection.

In comparison with other low-cost tests, the overall sensitivity and specificity to detect phenotypes for HbS and β-thalassemia were (ClinicalTrials.gov Identifier: NCT05506358): Gazelle Hb variant test (97.0% & 99.3%), HemoTypeSC (74.4% & 94.4%), and Sickle SCAN (75.0% & 94.7%)^37,38^. In terms of cost, the other point-of-care techniques are 2-10 times more expensive than the augmented sickling test for running 10,000-100,000 tests (Fig. 7), where the costs for the augmented sickling test include a one-time cost for the automated microscope (< $3,000), and inexpensive readily-available consumables (<$0.5 per test) such as microscope slides, coverslips, powdered reagent, and distilled water. Furthermore, only the Gazelle and the augmented sickling test can detect β-thalassemia trait, which is critical to screen for in regions with high prevalence of both HbS and β-thalassemia.

**Fig. 7.**
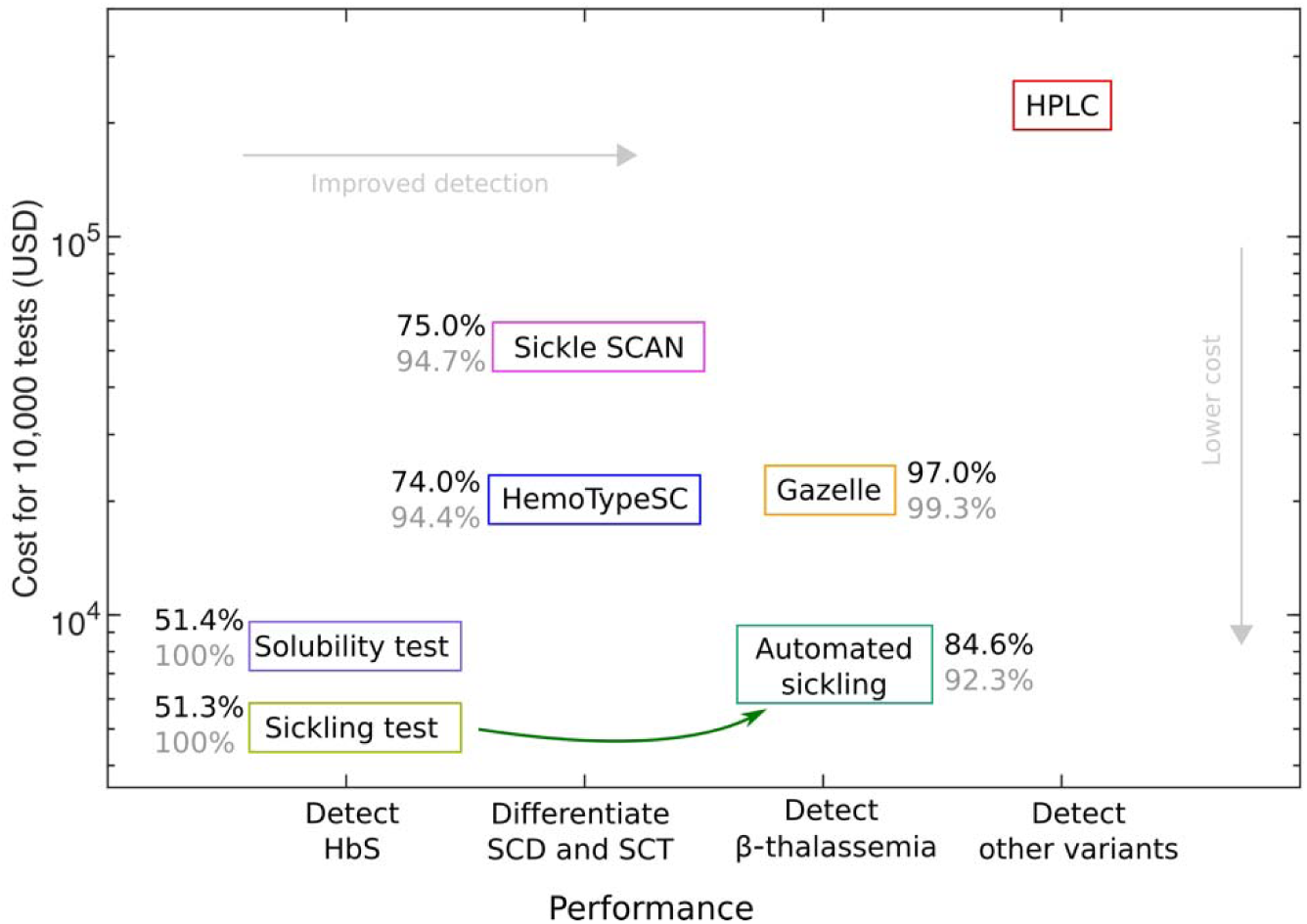
Cost performance plot for different detection techniques. Comparison of different low-cost techniques, including conventional sickling test, solubility test, Sickle SCAN, HemoTypeSC, Gazelle Hb variant test, and our augmented or automated sickling test, against the reference test (Hb HPLC). Numbers indicate sensitivity (black) and specificity (grey) for low-cost techniques against reference test (Hb HPLC). Numbers for sensitivity and specificity for the automated sickling test is for detecting 3 groups (*3GpSc*) for screening HbS and β-thalassemia: normal (HbAA), triat (HbA/β-thalassemia & AS), and SCD (HbSS & HbS/β-thalassemia).

There are some limitations associated with the diagnostic technique and with the data used in this study. Since the analysis relied on microscopic images, the morphological characterization was based on two-dimensional (2D) snapshots of three-dimensional (3D) cell morphologies. Although most round/healthy RBCs were flat and detected as round cells, some of these RBCs were on their sides, resulting in round cells being detected as elliptocytic cells, a common limitation of blood cell imaging-based diagnostic techniques. However, the occurrence of side-lying cells appeared to be negligible compared to flat-lying RBCs, and such cells appeared in both training and testing datasets. At the chosen configurations for time (2 hours) and temperature (room temperature), only a fraction of RBCs from SCT samples sickled. Although this served as a basis for differentiating between SCT and SCD, and resulted in high sensitivity to detect SCD, the incubation time or temperature can be increased to increase the number of sickled cells for SCT, likely increasing the sensitivity to detect SCT. Furthermore, some misclassifications between normal and β-thalassemia trait could possibly arise due to other factors such as iron deficiency (not evaluated in this study), which can affect the RBC shape and size. Future studies could include information such as ferritin level to account for other such confounding factors (*e.g.* iron deficiency) that affect cell morphology. The duration of storage for blood samples also affects RBC shape, and storage artifacts (some RBCs becoming echinocytic) were observed in some cells, which could limit the classification performance, especially between normal and β-thalassemia trait samples. Our study did not include β-thalassemia major patients because of their requirement for regular (*i.e.* around monthly) blood transfusions, which would affect the test results. Additionally, as HbC mutation is not encountered in Nepal, people with HbSC or HbAC were not included, and the algorithm therefore was not trained to distinguish these conditions.

In summary, automated microscopy and morphology-based classification enhanced the performance of the sickling test, and appears suitable to serve as a low-cost automated screening tool to detect sickle cell disease and β-thalassemia in low-resource settings, where the disease burden is often the highest.

## Materials and Methods

### Study design and participants

The study (ClinicalTrials.gov Identifier: NCT05506358) was conducted in two countries – Canada and Nepal. Blood collection and tests completed in Canada between September 2022 – March 2023 (at St. Paul’s Hospital and BC Children’s Hospital, Vancouver), and in Nepal in November and December 2022 (at Mount Sagarmatha Polyclinic and Diagnostic Center, Nepalgunj). A total of 138 participants were recruited (ages 2-74 years; 59% female, 41% male). The diagnoses for all participants, except normal controls or healthy volunteers without known β-globin disorders, had been previously established by hemoglobin high performance liquid chromatography (Hb HPLC). Confirmatory Hb HPLC was performed on all participants and there were 30 normal (HbAA), 23 β-thalassemia trait (HbA/β-thalassemia), 45 sickle cell trait (HbAS), and 40 sickle cell disease (11 HbS/β-thalassemia and 29 HbSS) participants.

Participants older than 1 year were eligible. Exclusion criteria were pregnancy or blood transfusion within 3 months. Informed consent was provided by the participants or parents, according to protocols approved by institutional/national research ethics boards. The study protocols and documents were reviewed and approved for the Canada study by the University of British Columbia-Providence Health Care Research Institute (UBC-PHC REB Number: H21-01929), and for the Nepal study by the University of British Columbia Clinical Research Ethics Board (UBC CREB Number: H22-00294) and Nepal Health Research Council (NHRC Registration Number: 85/2022). Additionally, institutional certificates of approval were obtained from the respective clinical sites where blood samples were stored and tested (from UBC Children’s & Women’s Research Ethics Board and from Mount Sagarmatha Polyclinic and Diagnostic Center).

### Blood sample collection

Blood samples were collected by trained phlebotomists or clinical laboratory technologists from veins in the antecubital fossa or the dorsum of the hand. In Canada, blood was drawn from blood collection needles or butterfly needles directly into 4.0 mL vacuum blood collection tubes with K2 EDTA (BD Vacutainer, Becton, Dickinson and Company). In Nepal, blood was drawn from hypodermic needles into syringes, and immediately transferred to 3.0 mL disposable non-vacuum blood collection tubes with K3 EDTA (AV LabotUbe, AV Consumables). Blood samples were stored in a refrigerator at 4°C. Before running any tests, blood samples were placed at room temperature for at least 30 minutes, and the blood tubes were repeatedly inverted or placed in rotary mixers to gently mix the separated plasma and settled blood cells. The blood samples were de-identified and associated with unique participant codes, which were used as references for the blood tests and analyses. For the automated sickling test, samples were prepared and imaged 1.4 ± 0.9 days (mean ± standard deviation) after blood collection, and the maximum time between blood collection and imaging was 2 days in Nepal and 4 days in Canada.

### Sample preparation for automated sickling test

The sickling test is a well-established and commonly used screening technique to detect the presence of hemoglobin S (HbS), but is not traditionally used to distinguish between sickle cell trait (SCT) and sickle cell disease (SCD)^7,13^. In our sample preparation method for the sickling test, red blood cells formed a monolayer with minimum overlap and adequate spacing for high-throughput imaging and subsequent image processing.

The sample preparation included the following steps:

1. Blood samples stored in EDTA tubes were taken out of storage (at 4°C) and placed at room temperature for at least 30 minutes.
2. The reagent, 2% (w/w) sodium metabisulphite mixed in distilled water, was freshly prepared and used up to around 3 hours after preparation. The reagent helped create a hypoxic environment, which facilitated sickling.
3. The separated plasma and settled blood cells were gently mixed by inverting the tube or placing it in a rotary mixer.
4. Whole blood and the reagent were pipetted at a 1:2 volume ratio (*e.g.* 150 µL of whole blood: 300 µL of reagent) into a separate tube or vial, and mixed by gentle inversions or in a rotary mixer.
5. A drop of the mixture of blood and reagent (3 µL to 3.3 µL) was pipetted onto a glass microscope slide, (VWR Vistavision 3” x 1” x 1 mm Microscope Slides, Cat. No. 16004-430).
6. A microscope coverslip (Fisherbrand Microscope Cover glass, 18 mm x 18 mm x 1 mm, 12542A, Fisher Scientific, Pittsburgh, PA, USA) was gently placed on top of the drop of blood and reagent to spread the mixture uniformly. The coverslip was gently lowered onto the mixture with the aid of a toothpick to spread the mixture while minimizing the formation of bubbles. The size of the coverslip (18 mm x 18 mm) was sufficient to spread the selected volume of the mixture (around 3 µL) uniformly, forming a monolayer of cells and with minimal leakage from the coverslip edges. A coverslip of a different size requires a different volume of the mixture to be dispensed.
7. The edges of the coverslip were sealed with a mixture of Vaseline, Lanolin and Parrafin (VALAP) at a 1:1:1 mass ratio^39^. The VALAP was melted in a glass petri dish placed on an electric hot plate at a temperature between 100°C and 110°C, and a paint brush was used to collect the melted VALAP and spread it on the edges of the coverslip. The VALAP solidified within a few seconds on the coverslip and microscope slide to seal the mixture of blood and reagent.
8. The process was duplicated for each slide for the same blood sample, such that there were two coverslips on each slide.

The sample preparation method for the sickling test from previous studies^7,13^ was modified to make it suitable for imaging. Firstly, larger volumes of blood and reagent (150 µL: 300 µL) were mixed in a tube or a vial, instead of mixing a small drop of blood with a small drop of reagent on the glass slide. Mixing the contents in a tube was more consistent than mixing them on the glass slide. Secondly, the volume ratio of blood to reagent was chosen as 1:2 (instead of the original 1:1 ratio) to avoid crowded cells and rouleaux formation. Additionally, an inert and non-toxic sealant, VALAP, was used instead of nail lacquers, which can contaminate the sample and sometimes created ghost cells (cells that have low contrast in images, due to negligible hemoglobin content and ruptured cell membranes^32^) at the periphery of the coverslip. Lastly, imaging was performed at 2 hours after sample preparation, while the samples were kept at room temperature (20-25°C), which ensured that most of the cells for individuals with SCD sickled. We found that temperature affects the rate of sickling – increasing the temperature increased the rate of sickling (Figures S2-S8 in Supplementary information). Imaging could potentially be done earlier than 2 hours (*e.g.* 30 minutes to an hour) if samples were incubated at a higher temperature (*e.g.* 37 °C), but temperature incubation was not chosen for this study due to the added complexity and requirement for a well-functioning incubator, which may not be accessible in remote/rural or low-resource settings.

### Automated high-throughput microscopy using Octopi

The sealed wet preparation of blood mixed with 2% sodium metabisulphite was imaged with a low-cost automated microscopy platform, Octopi (Open configurable high-throughput imaging platform for infectious disease diagnosis in the field)^26^. An infinity corrected 20× magnification objective lens (Olympus UPLFLN 20x/0.5) was connected through a tube lens to a monochromatic camera (MER2-1220-32U3M, Daheng Imaging). The monochromatic camera had a higher resolution than that of an equivalent color camera, and was sufficient (pixel size: 1.85μm) for characterizing the morphology of unstained cells in the prepared blood films. The microscope assembly included a 60 mm × 60 mm motorized XY translational stage (HDS-U-XY6060SN, Heidstar Co., Ltd.) for automated slide scanning, and a piezoelectric actuator to move the optical assembly for automated focusing. An LED matrix array illuminated the sample, and the illumination intensity, timing and number of active LEDs were programmatically controlled. All the functionality of the microscope, such as the computational illumination, image capture, motorized slide translation and autofocusing, were controlled using a python-based graphical user interface (GUI), operated in Linux.

The field of view (FOV) of each image was 3,000 pixels by 3,000 pixels, corresponding to 0.9 mm by 0.9 mm. At each sample location, an image pair was captured, by sequentially illuminating the left and right halves of the LED matrix for the individual images, so that the image pair was combined into a higher contrast image (discussed later in ‘DPC image processing’). For each coverslip, a 15 by 15 grid of image pairs or 225 image pairs were captured covering most of the area of the coverslip (with no overlap between adjacent FOVs), which took roughly 4 minutes including regular autofocusing after every 3 image pairs. For each participant, 6 coverslips (3 slides prepared in Nepal) or 12 coverslips (6 slides prepared in Canada) were imaged 2 hours after sample preparation. Some coverslips were imaged right after sample preparation, and at different time intervals (*e.g.* at configurations such as 1 hour, 3 hours, 4 hours, *etc.*). Time series imaging was also captured for some participants (*e.g.* 1000 image pairs captured every 3 seconds) to visualize the sickling process of red blood cells with different HbS concentrations. The images captured 2 hours after sample preparation were used for statistical analysis and machine learning classification discussed here.

### Reference or gold standard test

Hemoglobin high performance liquid chromatography (Hb HPLC) is one of the standard clinical methods used for detecting sickle cell disease, β-thalassemia and other common hemoglobinopathies, and was the reference test in Nepal and Canada^7^. All the blood samples were tested using the Hb HPLC technique; the D10 Hemoglobin Testing System (Bio-Rad Laboratories Inc., California, USA) was used in Nepal, and the BioRad Variant II (Bio-Rad Laboratories Inc., California, USA) was used in Canada. The interpretations of the Hb HPLC results by medical laboratory technologists or hematopathologists served as ground truth or diagnosis for the participants and was used for morphological characterization and classification.

### Differential phase contrast (DPC) image processing

The raw images from Octopi included image pairs illuminated from opposite illumination angles – one image illuminated by the left half (with intensity I_L_), and the other image illuminated by the right half (with intensity I_R_), of the programmable LED matrix. By normalizing the difference between the two images, the intensity of differential phase contrast (DPC) image I_DPC_ = 0.5 + (I_L_ - I_R_)/(I_L_ + I_R_) was computed on a pixel-by-pixel basis. DPC imaging captures the phase gradient information from the two images with opposite illumination angles, and does not rely on specialized objectives for phase imaging^40^. The blood cells in DPC images had sufficient contrast for morphological characterization (as shown in Fig. 1), without the need for staining, which is typically required for peripheral blood smears^32^. The DPC processing and most of the following image analyses were performed on a computing cluster (Cedar, Compute Canada, Digital Research Alliance of Canada) due to the large scale of the image dataset, which benefitted from computational resources and parallel computing on the cluster (mostly using GPUs).

### Segmentation of blood cells using Cellpose 2.0

Blood cells in the differential phase contrast (DPC) images were segmented using a generalist segmentation algorithm, Cellpose 2.0^30^, where a pre-trained neural network model (Cyto) was fine-tuned using a human-in-the-loop approach. The human-in-the-loop approach used 125 image segments (1,500 pixels × 1,500 pixels; 450 µm × 450 µm) from multiple donors and a recursive training approach. Each image in the training cycle required user annotations for correcting existing outlines predicted by the neural network and adding missing outlines of cells. The human-in-the-loop or iterative retraining approach greatly reduces the number of user annotations required because annotations are corrected in each retraining cycle, which iteratively improves the neural network model^30^. The default pre-trained neural-network model, Cyto, in Cellpose 2.0 already segmented round cells well, but the human-in-the-loop approach was necessary to segment sickle cells, to avoid segmenting platelets, artifacts (*e.g.* bubbles), and incomplete or obscured cells (*e.g.* overlapped cells and cells at the edges of the image).

Cell segmentation was the most computationally expensive process out of all the image processing tasks, taking around 1-2 minutes for segmentation of around 4,000-6,500 cells per image using GPUs. However, the overall computation time was significantly reduced by running parallel jobs on the Cedar cluster.

### Calculating morphological and intensity parameters

The outlines of segmented cells were saved by Cellpose 2.0 as text files, which were read by an image processing software, ImageJ^41^ (or FIJI^42^). The outlines were overlaid on the corresponding DPC images in ImageJ to calculate morphological and intensity parameters of each cell. The parameters were measured using ImageJ on the Cedar cluster, without the GUI or in “headless” mode. A total of 32 different morphological and intensity parameters were extracted for each cell, including basic morphological parameters (*e.g.* area, perimeter, minor axis, major axis, etc.), non-dimensional morphological parameters (*e.g.* circularity, aspect ratio, eccentricity, etc.), and intensity-based parameters (*e.g.* mean intensity, skewness, kurtosis, etc.). Furthermore, 8 basic morphological parameters were normalized by the mean values per image (*e.g.* normalized area, normalized perimeter, *etc.*). A complete list of these parameters is provided in Supplementary information. The data for each image (morphological and intensity parameters for every cell in the image) was stored in a comma separated variable or .csv files, which was used for further processing (*e.g.* using MATLAB programs).

### Morphological characterization

Normalized frequency distribution of each morphological and intensity parameter was calculated using the *histcounts* function in MATLAB, where the value in each bin v_l_ = c_l_ /N represented the ratio of the number of elements in the bin, c_l_, to the total number of elements in the data, N. The frequency distribution, representing relative probability of each morphological parameter, was calculated at different levels or scales – at the image level, the coverslip level, the participant or donor level, and the group or class level. For the analysis presented here, each frequency distribution was divided into 30 bins. The data for Canada and Nepal, imaged at 2 hours after sample preparation and stored at room temperature, were combined to form 4 clinically relevant groups – AA, ABeta, AS, and SCD (including SBeta and SS), and classification was performed on different combinations of these groups.

### Calculating statistical differences between different classes

Differences between the frequency distributions of the 4 different groups were calculated in MATLAB using one-way analysis of variance (ANOVA) and a post-hoc multiple comparison test (MCT), using functions *anova1* and *multcompare*. ANOVA compares the means of several groups and tests the hypothesis that they are equal against the alternative that they are not, indicating that at least one of the means is different without specifying which pairs are different. MCT can be applied for a more detailed pairwise comparison of the groups, by testing all the different combinations (6 combinations for 4 groups). The most conservative method for multiple comparison, the Scheffe’s method, was applied as it is suitable for exploratory data analysis^43^. MCT with Scheffe’s method was used to calculate p-values for all pair-wise combinations of the 4 groups at each bin of the frequency distribution of each morphological parameter. The bins or regions of all non-dimensional morphological parameters that resulted in the lowest p-values (highest differences) for each combination and for all combinations (considering the geometric mean of p-values) were highlighted and also selected for classification with feature selection.

### Morphology-based machine learning classification

The frequency distribution at the coverslip level (where each coverslip had around 225 images), describing the frequency of a parameter for all the cells under a coverslip for a particular participant, was used for classification of the different groups. Based on statistical analysis and due to the robustness of non-dimensional parameters, 19 non-dimensional morphological parameters were selected out of the total 40 parameters, as described in Supplementary information. The DPC images were not directly used for classification, but the morphological parameters were used instead because classification using the frequency distribution of these parameters was computationally much less resource-intensive and was also more interpretable than image-based classification – for instance, the frequency distribution of morphological parameters for classification can be visualized and compared statistically to check for differences between different classes.

Different groups were considered for classification: i) 3 groups (AA & ABeta, AS, SCD) typically considered for screening only HbS without β-thalassemia (referred to as 3 groups or *3Gp*). In this case, β-thalassemia trait (ABeta) was not identified separately from normal (AA), thus *3Gp* classification is suitable either in regions with low prevalence of β-thalassemia, or in regions with high β-thalassemia prevalence with supplemental low-cost screening tests for β-thalassemia, *e.g.* NESTROFT^44^. ii) 3 groups (AA, ABeta & AS, SCD) relevant for screening for both HbS and β-thalassemia, which combined the trait conditions together and the SCD conditions together (referred to as 3 groups for screening or *3GpSc*). The trait conditions, β-thalassemia trait (ABeta) and sickle cell trait (AS), were combined in *3GpSc* because of similar outcomes for the two asymptomatic groups after screening, *i.e.* follow-up confirmatory tests and genetic counselling for participants of reproductive age. iii) 4 clinically relevant groups (AA, ABeta, AS, SCD), where only SBeta and SS were combined because of their similar morphologies and clinical outcomes^3,5,7^ (referred to as 4 groups or *4Gp*).

The Classification Learner App in MATLAB (using MATLAB 2023a) was used to test the performance of 30 different models or classifiers (based on support vector machine, decision tree, ensemble, neural network, k-nearest neighbors, Naïve Bayes, and discriminant analysis – complete list in Supplementary information). An 80:20 participant-wise split of training and testing data was used such that data from each participant was either in the training split or the testing split, to prevent bias. The different classes were balanced by up-sampling or randomly repeating some data for the minority classes, such that all the classes had an equal number of images. Balancing the classes was important to remove bias towards the majority class when training the models and when calculating performance metrics. During training, 10-fold cross-validation was performed to reduce overfitting of the training dataset. As an example for the numbers of coverslips used for training and testing datasets, for the top classifier (subspace discriminant) for classification into 4 groups, the mean (and standard deviation) numbers for training (pre-balancing) was 798.8 (20.2) for all groups, 169.7 (6.0) for AA, 108.8 (0.4) for ABeta, 266.4 (6.4) for AS, and 253.9 (7.4) for SCD, and for testing (pre-balancing) was 202.2 (20.2) for all groups, 42.3 (6.0) for AA, 30.2 (0.4) for ABeta, 66.6 (6.4) for AS, and 63.1 (7.4) for SCD.

The performance metrics for validation (*e.g.* validation accuracy) obtained during training are not presented here, and only the performance metrics for testing (*e.g.* testing accuracy) are presented. In general, validation accuracy was higher than testing accuracy.

### Performance evaluation metrics

The sensitivity (or recall), Sens = TP/(TP + FN), is related to true positive (TP) and false negative (FN) values, and is the ability to correctly classify an individual belonging to a certain class or group. The specificity, Spec = TN/(TN + FP), is related to true negative (TN) and false positive (FP) values, and is the ability to correctly classify an individual not belonging to a certain class or group. Other performance metrics, such as accuracy, positive predictive value (or precision), negative predictive value, and F1-score, were also calculated and are tabulated for the top 5 classifiers in Supplementary information. For multiclass problems, the performance metrics can be calculated, per class or group, by comparing one group against all others (also known as one vs. all or one vs. rest), and the metrics were macro-averaged by taking the mean of values for all individual groups.

### Classification metrics, repeatability and confidence intervals

To test the repeatability of the classifications and to obtain confidence intervals for the evaluation metrics, all classification models were iteratively trained and tested using random participant-wise splits of the data (80: 20 for training: testing) for 1,000 independent iterations. For each iteration, all evaluation metrics were calculated for the testing data set: accuracy, sensitivity or recall, specificity, positive predictive value or precision, negative predictive value, and F1-score calculated from the confusion matrix, and the area under the curve (AUC) calculated from the receiver operating characteristic (ROC) curve for each class (using one vs. all method; Fig. 5). Macro-averaged values of these evaluation metrics were calculated to combine metrics from all classes. For all 1,000 iterations, the mean and 95% confidence intervals (assuming normal distribution) were calculated for each evaluation metric and the macro-averaged metrics. Additionally, data from all 1,000 iterations were aggregated to calculate merged confusion matrices and ROC curves (individual class and macro-averaged ROC curves).

### Open-access database

In accordance with the approved research protocols and informed consent from participants, de-identified data (such as de-identified images of blood films from Octopi, and processed data such as segmentation outlines, morphological parameters, *etc.*) were deposited in an online public repository, Federated Research Data Repository (FRDR), an open-access repository hosted by the Digital Research Alliance of Canada. The link to the open-access dataset is provided https://doi.org/10.20383/103.091631.

The dataset contains all the raw and processed data, and can be used to reproduce the results presented here and also for further development of classification or morphological characterization of sickle cell disease and β-thalassemia. The morphological and intensity parameters for data from Nepal and Canada used in the current work (imaging at 2 hours at room temperature), and additional data for additional configurations (*e.g.* different time settings, temperature settings, and time series data) are available publicly in FRDR, stored as .csv files.

All the associated code (written in Python, MATLAB, or shell) for generating results presented here are also available online in a public Github repository (https://github.com/p-shrestha/erythroSight).

## Data Availability

In accordance with the approved research protocols and informed consent from participants, de-identified data (such as de-identified images of blood films from Octopi, and processed data such as segmentation outlines, morphological parameters, etc.) were deposited in an online public repository, Federated Research Data Repository (FRDR), an open-access repository hosted by the Digital Research Alliance of Canada. The link to the open-access dataset is provided https://doi.org/10.20383/103.0916.
All the associated code (written in Python, MATLAB, or shell) for generating results presented here are also available online in a public Github repository (https://github.com/p-shrestha/erythroSight).

## Acknowledgements

We sincerely thank all the participants in the study for their invaluable blood samples; Ashik Gurung, Mahesh Chaudhary, Ram Prabesh Tharu, Asmita Chaudhary, Sanjeev Chaudhary, and Shiva Gautam for blood collection, tests and HPLC interpretation at Mount Sagarmatha Polyclinic and Diagnostic Center in Nepal; Dr. Mykola Maydan for help with coordination, equipment setup and technical advice at the pathology lab at BC Children’s Hospital; all the phlebotomists and staff at BC Children’s Hospital and St. Paul’s Hospital in Canada who helped with the study; Dinesh Raj Sapkota and Nura Basnet from Creating Possibilities Nepal, and the UBC sickle cell team for helpful discussions; Dr. Roshan Chitrakar for document translations; Erin Clary, Eugene Barsky, Jiarui Li, Paul Lesack, Tamanna Moharana, and Nick Rochlin for helping set up the open-access dataset, cloud computing tools and data transfer protocols. This research was undertaken, in part, with support from the Canada Research Chairs program, UBC Health Innovation Funding Investment (HIFI) Awards, and the UBC Four Year Doctoral Fellowship (4YF) program. The UBC Centre for Blood Research is home to the Naiman Vickars Endowment fund which has provided funds for this project.

## Author contributions

Pranav Shrestha: Conceptualization, Methodology, Software, Formal analysis, Investigation, Writing - Original Draft, Writing - Review & Editing, Visualization, Funding acquisition. Hendrik Lohse: Software, Investigation, Writing - Review & Editing. Christopher Bhatla: Conceptualization, Methodology, Investigation, Writing - Review & Editing. Heather McCartney: Resources, Project administration. Alaa Alzaki: Resources. Navdeep Sandhu: Resources, Project administration. Pradip Kumar Oli: Investigation, Project administration. Hongquan Li: Methodology, Software, Resources, Writing - Review & Editing. Manu Prakash: Methodology, Resources, Supervision, Writing - Review & Editing. Ali Amid: Methodology, Resources, Writing - Review & Editing. Rodrigo Onell: Conceptualization, Methodology, Writing - Review & Editing. Nicholas Au: Conceptualization, Methodology, Writing - Review & Editing. Hayley Merkeley: Conceptualization, Methodology, Resources, Writing - Review & Editing. Videsh Kapoor: Conceptualization, Methodology, Writing - Review & Editing, Funding acquisition. Rajan Pande: Conceptualization, Methodology, Resources, Supervision. Boris Stoeber: Conceptualization, Methodology, Formal analysis, Writing - Original Draft, Writing - Review & Editing, Supervision, Funding acquisition.

## Competing interests

Hongquan Li and Manu Prakash are co-founders, and Pranav Shrestha is an employee, of Cephla Inc., which is a spin-out company from Stanford University, deploying Octopi globally for disease diagnostics.

## Notes

### Clinical Trial

NCT05506358

### Author Declarations

Ethics committee/IRB University of British Columbia-Providence Health Care Research Institute (UBC-PHC REB Number: H21-01929) of the University of British Columbia gave ethical approval for this work (for the study in Canada) Ethics committee/IRB University of British Columbia Clinical Research Ethics Board (UBC CREB Number: H22-00294) of the University of British Columbia, and Nepal Health Research Council (NHRC Registration Number: 85/2022) of Nepal (national research ethics review board) gave ethical approval for this work (for the study in Nepal)

