## Supplementary information for "Morphology-based classification of sickle cell disease and β-thalassemia using a low-cost automated microscope and machine learning"

[5]Division of Hematology, Providence Health Care, 440-1144 Burrard Street, Vancouver, British Columbia, V6Z 2A5, Canada.

[6]Mount Sagarmatha Polyclinic and Diagnostic Center, Nepalgunj, Bheri Zone, Province no-5, Nepal.

[7]Department of Electrical Engineering, Stanford University, 350 Jane Stanford Way, Stanford, California, USA.

[8]Department of Bioengineering, Stanford University, 443 Via Ortega, Stanford, California, USA.

[9]Department of Internal Medicine, Bheri Hospital, Nepalgunj, Bheri Zone, Province no-5, Nepal.

[10]Department of Electrical and Computer Engineering, The University of British Columbia, 2332 Main Mall, Vancouver, British Columbia, V6T 1Z4, Canada.

### List of supplementary videos

Supplementary video 1: Example of sickling at the cellular level for a sample with sickle cell disease. Video playback speed is 20× real-time.

Supplementary video 2: Example of sickling at the cellular level for a sample with sickle cell trait. Video playback speed is 20× real-time.

Supplementary video 3: Example of sickling at the image level (same as Fig 2b-g), showing differential phase contrast image of red blood cells and morphological characterization of roundness and eccentricity. Cells are predominantly round at t = 0 s, and change to sickled shapes by 3000 s. Number of cells = 2805 ± 17 (mean ± standard deviation for 1000 images). Video playback speed is 120× real-time.

### Automated microscope

The automated microscope, Octopi, used for both studies in Canada and Nepal, is shown in Fig. S1.

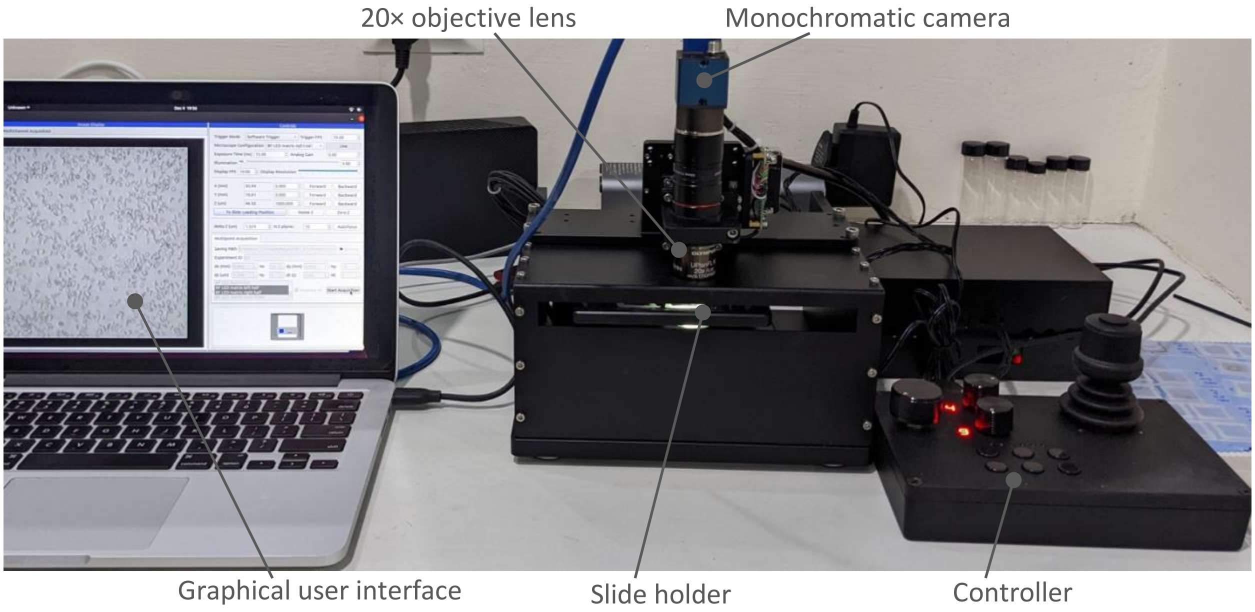


**Fig. S1**: Picture of the automated microscope, Octopi, during imaging of 225 images/coverslip at the study site in Nepal.

#### Morphological and intensity parameters

A complete list of 40 morphological and intensity-based parameters extracted for each cell is provided below, with the numbers corresponding to those in Fig. 4 i and j in the main text.

| Number | Morphological or intensity parameter | Type | Formula |
|---|---|---|---|
| 1 | Area | Basic morphological | |
| 2 | Perimeter | Basic morphological | |
| 3 | Angle | Basic morphological | |
| 4 | Major axis | Basic morphological | |
| 5 | Minor axis | Basic morphological | |
| 6 | Height | Basic morphological | |
| 7 | Width | Basic morphological | |
| 8 | Mean intensity | Intensity | |
| 9 | Standard deviation intensity | Intensity | |
| 10 | Median intensity | Intensity | |
| 11 | Skewness | Intensity | |
| 12 | Kurtosis | Intensity | |
| 13 | Feret | Basic morphological | |
| 14 | Feret angle | Basic morphological | |
| 15 | Minimum feret | Basic morphological | |
| 16 | Feret X coordinate | Basic morphological | |
| 17 | Feret Y coordinate | Basic morphological | |
| 18 | Circularity | Non-dimensional morphological | $\frac{4 \cdot \pi \cdot Area}{Perimeter^2}$ |
| 19 | Aspect ratio | Non-dimensional morphological | $\frac{Major}{Minor}$ |
| 20 | Roundness | Non-dimensional morphological | $\frac{4 \cdot Area}{\pi \cdot Major^2}$ |

| | | | |
|---|---|---|---|
| 21 | Convex hull area | Basic morphological | |
| 22 | Solidity | Non-dimensional morphological | $\frac{Area}{Convex\ Hull\ Area}$ |
| 23 | Eccentricity | Non-dimensional morphological | 1. $\sqrt{1-\frac{0.5\cdot Minor^2}{0.5\cdot Major^2}}$ |
| 24 | Elliptical shape factor (ESF) | Non-dimensional morphological | $\frac{1}{AR}=\frac{Minor}{Major}$ |
| 25 | ElongationFW | Non-dimensional morphological | $\frac{Width}{Feret}$ |
| 26 | ElongationFmF | Non-dimensional morphological | $\frac{Minimum\ Feret}{Feret}$ |
| 27 | Convex perimeter | Basic morphological | |
| 28 | Convexity | Non-dimensional morphological | $\frac{Convex\ Perimeter}{Perimeter}$ |
| 29 | Fiber length | Basic morphological | |
| 30 | Fiber width | Basic morphological | |
| 31 | Curl | Non-dimensional morphological | $\frac{Feret}{Fibre\ Length}$ $=\frac{4\cdot Feret}{Perimeter-\sqrt{Perimeter^2-16\cdot Area}}$ |
| 32 | CurlW | Non-dimensional morphological | $\frac{Feret}{Fibre\ Width}=\frac{Feret\cdot Fibre\ Length}{Area}$ |
| 33 | Normalized area | Normalized | |
| 34 | Normalized perimeter | Normalized | |
| 35 | Normalized major axis | Normalized | |
| 36 | Normalized minor axis | Normalized | |
| 37 | Normalized height | Normalized | |
| 38 | Normalized width | Normalized | |
| 39 | Normalized feret | Normalized | |
| 40 | Normalized minimum feret | Normalized | |

#### List of models or classifiers

The complete list of different models/classifiers in MATLAB Classification Learner App (2023a) is provided below:

1. Fine Tree
2. Medium Tree
3. Coarse Tree
4. Linear Discriminant
5. Quadratic Discriminant
6. Efficient Logistic Regression
7. Efficient Linear Support Vector Machine (SVM)
8. Gaussian Naive Bayes
9. Kernel Naive Bayes
10. Linear SVM
11. Quadratic SVM
12. Cubic SVM
13. Fine Gaussian SVM
14. Medium Gaussian SVM
15. Coarse Gaussian SVM
16. Fine K-Nearest Neighbors (KNN)
17. Medium KNN
18. Coarse KNN
19. Cosine KNN
20. Cubic KNN
21. Weighted KNN
22. Boosted Tree (Ensemble)
23. Bagged Tree (Ensemble)
24. Subspace Discriminant (Ensemble)
25. Subspace KNN (Ensemble)
26. RUS Boosted Tree (Ensemble)
27. Narrow Neural Network
28. Medium Neural Network
29. Wide Neural Network
30. Bi-layered Neural Network
31. Tri-layered Neural Network
32. SVM Kernel
33. Logisitic Regression Kernel

#### Performance metrics

Additional performance metrics F1-Score, accuracy (one-versus-all), positive predictive value (PPV) and negative predictive value (NPV) for the models described in the main text are provided in tables S1-S3.

**Table S1**: Additional performance metrics for 3 groups (AA & ABeta, AS, SCD) typically considered for screening only HbS without β-thalassemia (referred to as 3 groups or *3Gp*)

| | F1-Score | Accuracy (OvA) | PPV | NPV |
|---|---|---|---|---|
| **Quadratic SVM** | | | | |
| Macro-averaged | 0.889 (0.887-0.891) | 0.927 (0.925-0.928) | 0.893 (0.891-0.895) | 0.946 (0.945-0.947) |
| AA&ABeta | 0.862 (0.859-0.865) | 0.907 (0.905-0.909) | 0.852 (0.848-0.856) | 0.94 (0.938-0.942) |
| AS | 0.833 (0.829-0.836) | 0.891 (0.889-0.893) | 0.858 (0.853-0.862) | 0.91 (0.908-0.913) |
| SCD | 0.973 (0.971-0.974) | 0.982 (0.981-0.983) | 0.97 (0.968-0.972) | 0.989 (0.988-0.99) |
| **Linear SVM** | | | | |
| Macro-averaged | 0.881 (0.878-0.883) | 0.922 (0.921-0.924) | 0.894 (0.892-0.897) | 0.946 (0.945-0.947) |
| AA&ABeta | 0.864 (0.861-0.867) | 0.9 (0.898-0.902) | 0.794 (0.79-0.798) | 0.974 (0.973-0.976) |
| AS | 0.805 (0.8-0.809) | 0.884 (0.882-0.887) | 0.912 (0.908-0.916) | 0.877 (0.874-0.879) |
| SCD | 0.973 (0.972-0.975) | 0.982 (0.982-0.983) | 0.977 (0.975-0.979) | 0.986 (0.985-0.987) |
| **Wide Neural Net** | | | | |
| Macro-averaged | 0.882 (0.88-0.885) | 0.921 (0.92-0.923) | 0.888 (0.885-0.89) | 0.942 (0.941-0.943) |
| AA&ABeta | 0.862 (0.858-0.865) | 0.908 (0.906-0.91) | 0.861 (0.857-0.866) | 0.935 (0.933-0.937) |
| AS | 0.831 (0.828-0.835) | 0.886 (0.884-0.888) | 0.829 (0.825-0.833) | 0.92 (0.918-0.922) |
| SCD | 0.954 (0.951-0.956) | 0.97 (0.969-0.972) | 0.972 (0.97-0.974) | 0.971 (0.969-0.973) |
| **Cubic SVM** | | | | |
| Macro-averaged | 0.879 (0.877-0.882) | 0.92 (0.918-0.922) | 0.883 (0.881-0.886) | 0.941 (0.94-0.942) |
| AA&ABeta | 0.841 (0.838-0.845) | 0.895 (0.893-0.897) | 0.84 (0.836-0.845) | 0.926 (0.924-0.928) |
| AS | 0.822 (0.818-0.826) | 0.883 (0.88-0.885) | 0.836 (0.832-0.84) | 0.909 (0.907-0.912) |
| SCD | 0.974 (0.973-0.976) | 0.983 (0.982-0.984) | 0.973 (0.972-0.975) | 0.988 (0.987-0.99) |
| **Trilayered Neural Net** | | | | |
| Macro-averaged | 0.878 (0.876-0.88) | 0.919 (0.917-0.92) | 0.883 (0.881-0.885) | 0.94 (0.939-0.941) |
| AA&ABeta | 0.852 (0.849-0.855) | 0.902 (0.9-0.904) | 0.856 (0.851-0.86) | 0.929 (0.927-0.931) |
| AS | 0.825 (0.821-0.828) | 0.881 (0.879-0.884) | 0.823 (0.819-0.827) | 0.916 (0.914-0.918) |
| SCD | 0.958 (0.956-0.96) | 0.973 (0.971-0.974) | 0.971 (0.969-0.974) | 0.975 (0.973-0.977) |

OvA = one vs. all; PPV = positive predictive value; NPV = negative predictive value

**Table S2**: Additional performance metrics for 3 groups (AA, ABeta & AS, SCD) relevant for screening for both HbS and β-thalassemia, which combines the trait conditions together (referred to as 3 groups for screening or *3GpSc*)

| | F1-Score | Accuracy (OvA) | PPV | NPV |
|---|---|---|---|---|
| **Quadratic SVM** | | | | |
| Macro-averaged | 0.844 (0.84-0.847) | 0.897 (0.895-0.899) | 0.855 (0.853-0.858) | 0.926 (0.924-0.927) |
| AA | 0.764 (0.758-0.771) | 0.859 (0.856-0.862) | 0.839 (0.835-0.843) | 0.872 (0.868-0.875) |
| ABeta & AS | 0.79 (0.787-0.794) | 0.848 (0.844-0.851) | 0.747 (0.741-0.753) | 0.919 (0.917-0.921) |
| SCD | 0.976 (0.975-0.978) | 0.984 (0.984-0.985) | 0.981 (0.979-0.982) | 0.987 (0.986-0.988) |
| **Medium Gaussian SVM** | | | | |
| Macro-averaged | 0.83 (0.827-0.833) | 0.888 (0.885-0.89) | 0.837 (0.834-0.84) | 0.918 (0.916-0.919) |
| AA | 0.764 (0.758-0.77) | 0.844 (0.841-0.848) | 0.761 (0.757-0.765) | 0.893 (0.889-0.896) |
| ABeta & AS | 0.751 (0.747-0.755) | 0.835 (0.832-0.838) | 0.775 (0.768-0.782) | 0.873 (0.871-0.875) |
| SCD | 0.974 (0.973-0.976) | 0.983 (0.982-0.984) | 0.975 (0.973-0.976) | 0.988 (0.987-0.989) |
| **Cubic SVM** | | | | |
| Macro-averaged | 0.828 (0.824-0.831) | 0.887 (0.885-0.889) | 0.844 (0.841-0.847) | 0.919 (0.918-0.921) |
| AA | 0.728 (0.722-0.735) | 0.842 (0.839-0.845) | 0.832 (0.828-0.836) | 0.849 (0.846-0.853) |
| ABeta & AS | 0.775 (0.771-0.778) | 0.831 (0.828-0.834) | 0.712 (0.707-0.717) | 0.921 (0.919-0.923) |
| SCD | 0.98 (0.979-0.981) | 0.987 (0.986-0.988) | 0.988 (0.987-0.989) | 0.987 (0.986-0.988) |
| **Trilayered Neural Network** | | | | |
| Macro-averaged | 0.828 (0.825-0.832) | 0.886 (0.884-0.889) | 0.842 (0.839-0.845) | 0.918 (0.916-0.919) |
| AA | 0.751 (0.745-0.757) | 0.851 (0.848-0.854) | 0.826 (0.822-0.83) | 0.865 (0.861-0.868) |
| ABeta & AS | 0.771 (0.767-0.774) | 0.832 (0.829-0.835) | 0.721 (0.716-0.727) | 0.911 (0.909-0.913) |
| SCD | 0.963 (0.961-0.965) | 0.976 (0.975-0.978) | 0.978 (0.977-0.98) | 0.977 (0.975-0.978) |
| **Subspace Discriminant** | | | | |
| Macro-averaged | 0.828 (0.824-0.831) | 0.886 (0.884-0.888) | 0.835 (0.832-0.838) | 0.917 (0.915-0.918) |
| AA | 0.742 (0.736-0.748) | 0.837 (0.834-0.84) | 0.773 (0.768-0.777) | 0.871 (0.868-0.875) |
| ABeta & AS | 0.758 (0.754-0.762) | 0.833 (0.83-0.836) | 0.747 (0.741-0.753) | 0.888 (0.886-0.891) |
| SCD | 0.983 (0.981-0.984) | 0.989 (0.988-0.99) | 0.986 (0.985-0.988) | 0.99 (0.989-0.991) |

OvA = one vs. all; PPV = positive predictive value; NPV = negative predictive value

**Table S3**: Additional performance metrics for 4 groups (AA, ABeta, AS, SCD) referred as *4Gp*

| | F1-Score | Accuracy (OvA) | PPV | NPV |
|---|---|---|---|---|
| **Subspace Discriminant** | | | | |
| Macro-averaged | 0.762 (0.758-0.766) | 0.882 (0.88-0.883) | 0.777 (0.773-0.78) | 0.923 (0.921-0.924) |
| AA | 0.656 (0.65-0.663) | 0.822 (0.819-0.825) | 0.645 (0.638-0.652) | 0.895 (0.892-0.897) |
| ABeta | 0.651 (0.644-0.659) | 0.828 (0.824-0.831) | 0.66 (0.653-0.668) | 0.89 (0.887-0.893) |
| AS | 0.761 (0.756-0.766) | 0.887 (0.884-0.889) | 0.819 (0.812-0.825) | 0.912 (0.91-0.914) |
| SCD | 0.98 (0.979-0.982) | 0.99 (0.989-0.991) | 0.982 (0.981-0.984) | 0.993 (0.993-0.994) |
| **Quadratic SVM** | | | | |
| Macro-averaged | 0.746 (0.742-0.75) | 0.876 (0.874-0.878) | 0.757 (0.753-0.76) | 0.92 (0.919-0.921) |
| AA | 0.678 (0.673-0.684) | 0.836 (0.833-0.839) | 0.672 (0.666-0.678) | 0.9 (0.898-0.903) |
| ABeta | 0.579 (0.57-0.587) | 0.813 (0.81-0.816) | 0.655 (0.647-0.662) | 0.857 (0.854-0.86) |
| AS | 0.757 (0.752-0.761) | 0.87 (0.868-0.873) | 0.731 (0.725-0.738) | 0.931 (0.929-0.932) |
| SCD | 0.97 (0.968-0.972) | 0.985 (0.984-0.986) | 0.968 (0.966-0.97) | 0.991 (0.99-0.992) |
| **Linear SVM** | | | | |
| Macro-averaged | 0.75 (0.746-0.753) | 0.875 (0.873-0.877) | 0.769 (0.765-0.772) | 0.918 (0.917-0.92) |
| AA | 0.657 (0.651-0.663) | 0.812 (0.808-0.815) | 0.615 (0.608-0.622) | 0.904 (0.901-0.906) |
| ABeta | 0.612 (0.603-0.62) | 0.812 (0.808-0.815) | 0.627 (0.62-0.634) | 0.876 (0.873-0.88) |
| AS | 0.759 (0.754-0.764) | 0.891 (0.889-0.893) | 0.854 (0.848-0.86) | 0.905 (0.903-0.906) |
| SCD | 0.971 (0.97-0.973) | 0.986 (0.985-0.987) | 0.979 (0.978-0.981) | 0.989 (0.988-0.99) |
| **Wide Neural Network** | | | | |
| Macro-averaged | 0.733 (0.729-0.736) | 0.871 (0.869-0.872) | 0.747 (0.744-0.751) | 0.917 (0.916-0.918) |
| AA | 0.676 (0.67-0.682) | 0.83 (0.827-0.833) | 0.655 (0.649-0.662) | 0.903 (0.901-0.906) |
| ABeta | 0.536 (0.527-0.544) | 0.803 (0.8-0.805) | 0.647 (0.639-0.655) | 0.841 (0.839-0.844) |
| AS | 0.767 (0.763-0.772) | 0.873 (0.87-0.875) | 0.721 (0.715-0.726) | 0.942 (0.94-0.944) |
| SCD | 0.952 (0.95-0.955) | 0.977 (0.976-0.978) | 0.967 (0.964-0.969) | 0.981 (0.98-0.983) |
| **Boosted Tree** | | | | |
| Macro-averaged | 0.736 (0.732-0.74) | 0.87 (0.868-0.872) | 0.75 (0.746-0.754) | 0.915 (0.914-0.916) |
| AA | 0.68 (0.673-0.688) | 0.848 (0.845-0.851) | 0.723 (0.715-0.731) | 0.893 (0.89-0.896) |
| Abeta | 0.588 (0.58-0.596) | 0.797 (0.793-0.8) | 0.598 (0.591-0.606) | 0.869 (0.865-0.872) |
| AS | 0.73 (0.724-0.735) | 0.86 (0.857-0.863) | 0.721 (0.714-0.728) | 0.917 (0.915-0.919) |
| SCD | 0.948 (0.945-0.951) | 0.974 (0.973-0.975) | 0.959 (0.955-0.963) | 0.981 (0.98-0.982) |

#### Effect of temperature

Temperature increased the rate of sickling. This was more prominent in sickle cell trait samples than sickle cell disease samples (where majority of cells sickled even at room temperature), as shown in Fig. S2-S8.

##### Sickle cell disease

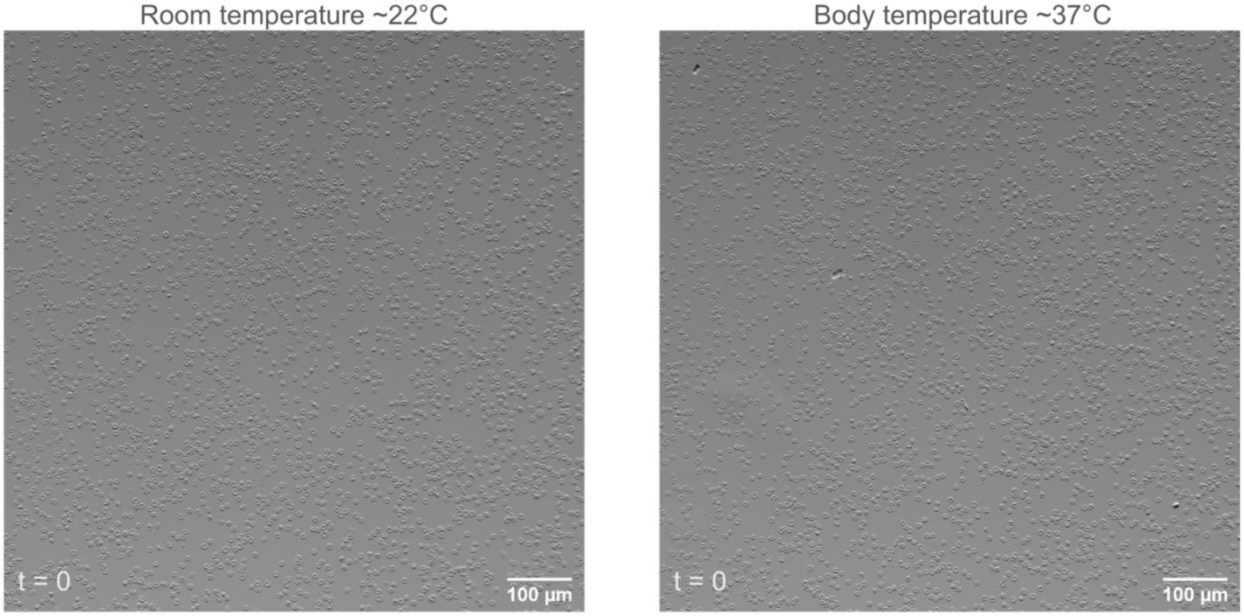


**Fig. S2**: Images of red blood cells from sickle cell disease sample at room temperature (left) and body temperature (right), taken immediately after sample preparation (t=0).

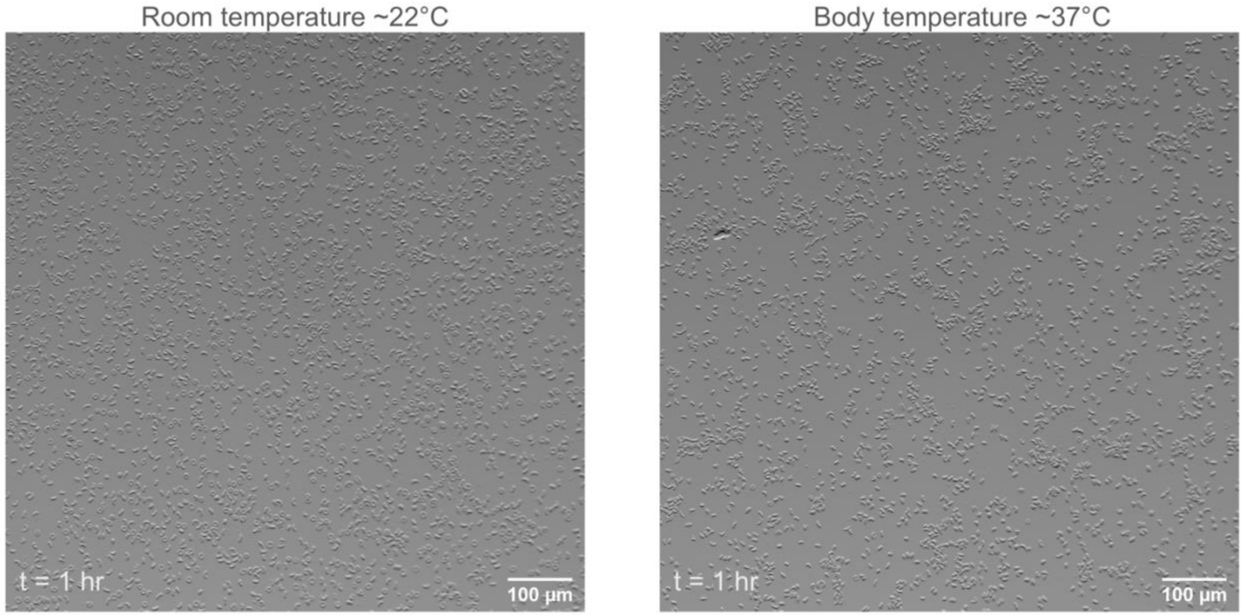


**Fig. S3**: Images of red blood cells from sickle cell disease sample at room temperature (left) and body temperature (right), taken 1 hour after sample preparation (t=1hr).

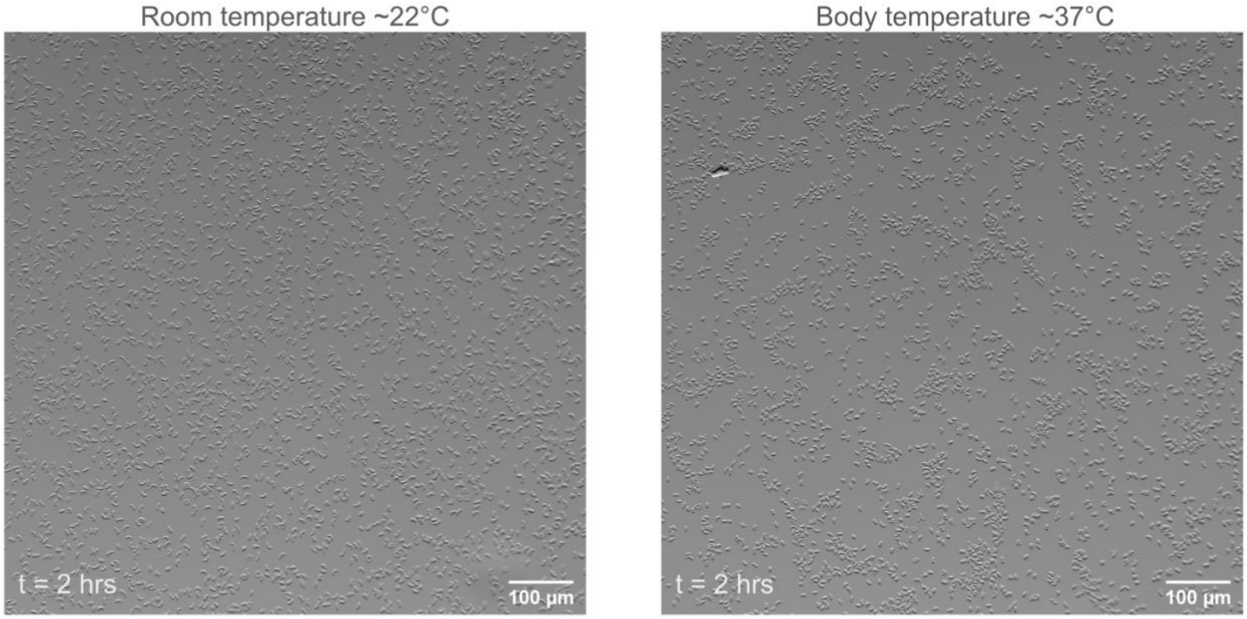


**Fig. S4**: Images of red blood cells from sickle cell disease sample at room temperature (left) and body temperature (right), taken 2 hours after sample preparation (t=2hrs).

#### Sickle cell trait

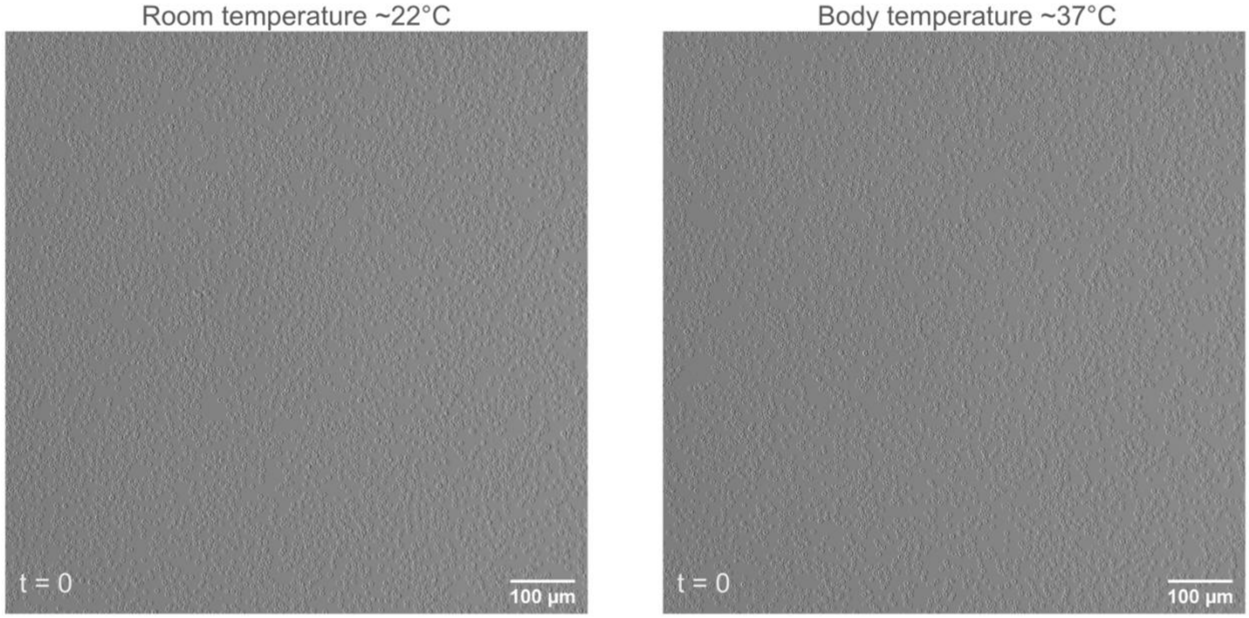


**Fig. S5**: Images of red blood cells from sickle cell trait sample at room temperature (left) and body temperature (right), taken immediately after sample preparation (t=0).

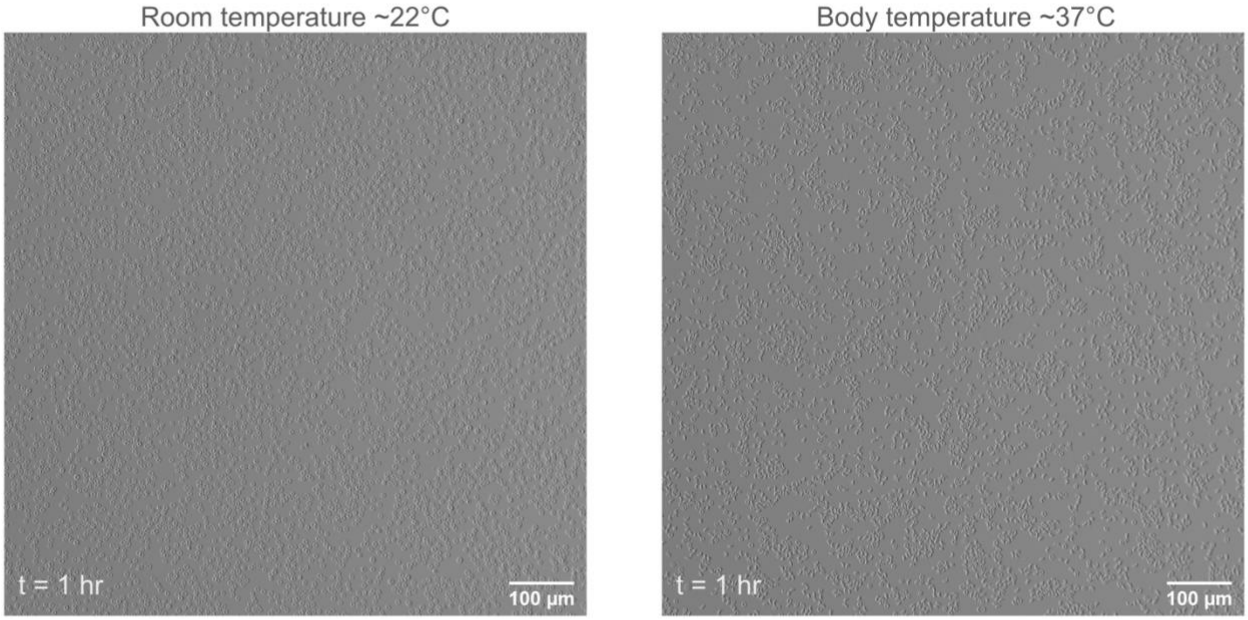


**Fig. S6**: Images of red blood cells from sickle cell trait sample at room temperature (left) and body temperature (right), taken 1 hour after sample preparation (t=1hr). Note: At room temperature there is negligible sickling, while at body temperature, most of the cells have sickled.

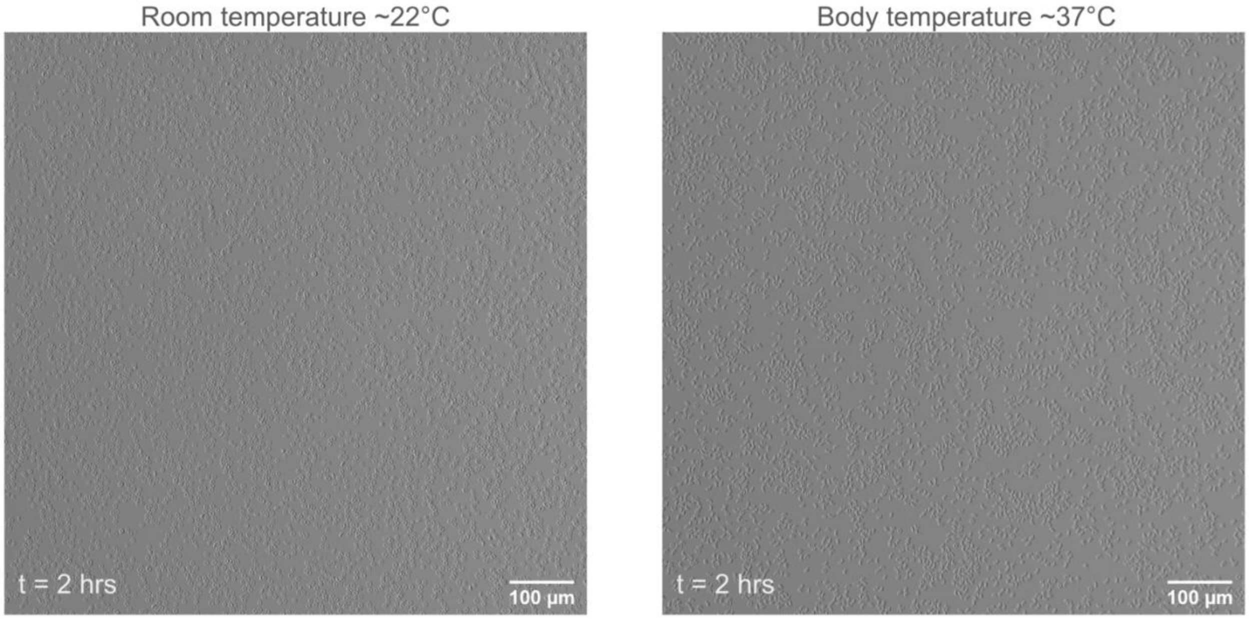


**Fig. S7**: Images of red blood cells from sickle cell trait sample at room temperature (left) and body temperature (right), taken 2 hours after sample preparation (t=2hrs). Note: At room temperature there is negligible sickling, while at body temperature, most of the cells have sickled.

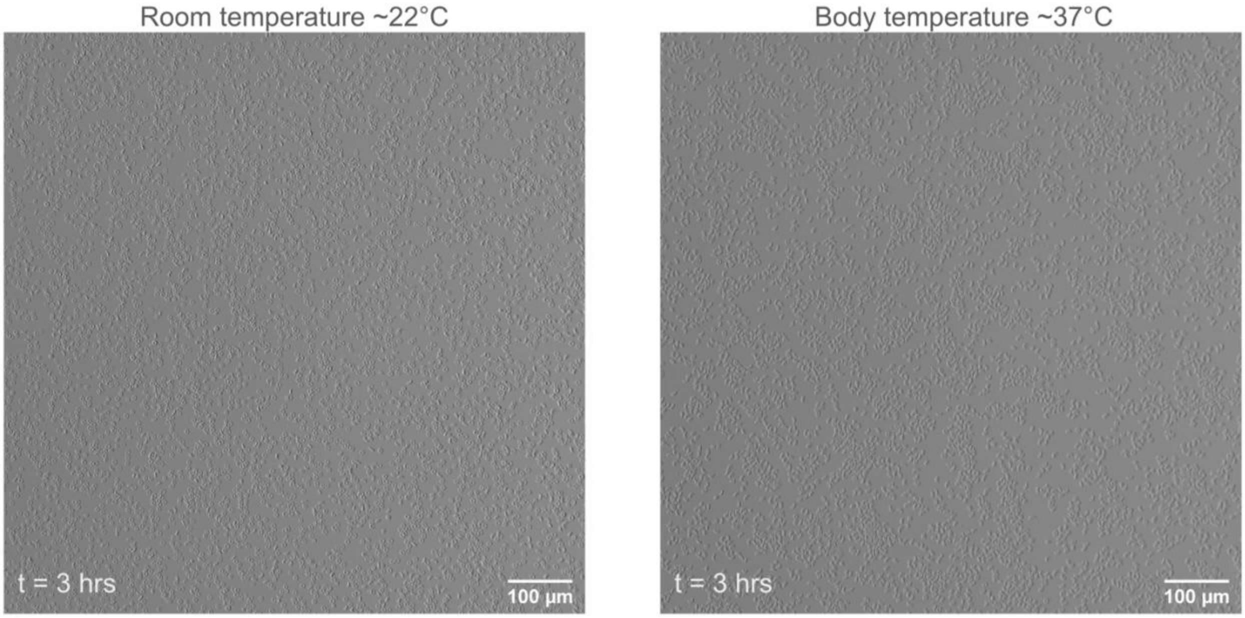


**Fig. S8**: Images of red blood cells from sickle cell trait sample at room temperature (left) and body temperature (right), taken 3 hours after sample preparation (t=3hrs).
